# Simulating Psychotherapeutic Sessions with Generative Artificial Intelligence: A Proof-of-Concept Study of *In Silico* Psychotherapy Research

**DOI:** 10.64898/2026.10.22.26366245

**Authors:** Katalin Horváth, Shiv Longani, Shir Brill, Harry A. Ballantyne, Rainer Schaefert, Marion Tegethoff, Liam Schirmer, Emma Börner, Ingo J. Timm, Zohar Elyoseph, Gunther Meinlschmidt

## Abstract

**Background:** Across biomedicine, *in silico* models increasingly complement *in vivo* and *in vitro* research. Psychotherapy science, however, remains constrained by methodological limitations, resource-intensive trials, challenges in manipulating clinically meaningful features experimentally, and a potential ceiling in efficacy. Leveraging generative AI and large language models (LLMs), we propose a framework for “*in silico* psychotherapy research”: controlled simulations of psychotherapeutic interactions under reproducible experimental conditions.

**Objective:** We test whether such simulation experiments can recover a theoretically expected effect of an isolated psychotherapy-process manipulation.

**Methods:** In a preregistered proof-of-concept study, we generated 512 fully automated motivational interviewing (MI) sessions by linking locally deployed Llama-3.3-70B-Instruct based therapist and patient models across eight vignettes and eight therapist personas. Therapist, but not patient, prompts manipulated common therapeutic factors (CTF), contrasting high versus low empathy, affirmation, and alliance building behaviour in a controlled design. The primary outcome was MI treatment integrity (MITI 4.2 global ratings), assessed by a blinded LLM with repeated ratings and analysed using linear mixed-effects models. Secondary outcomes included observer-inferred importance/confidence rulers based on simulated patient utterances; Therapist Empathy Scale (TES) and Working Alliance Inventory–Short Observer (WAI-S-O) served as manipulation fidelity checks. Human coders rated a stratified subsample.

**Results:** Of the 512 simulations, 506 (98.8%) yielded complete transcripts and 480 (93.75) were retained for analysis. Manipulation checks confirmed the fidelity of the CTF manipulation (WAI-S-O total scores β=3.14; [2.65,3.63], p<0.001, TES β=14.48 [13.92,15.04], p<0.001). High versus low CTF produced a theoretically expected improvement in all MITI global ratings, including the MI-specific features not directly targeted by the CTF manipulation (β=0.54–0.95; all p<0.001). Simulated patient confidence but not importance for change increased more under high CTF (β=0.17 [0.09,0.26], p<0.001). Human ratings confirmed the direction of the primary contrast.

**Conclusions:** LLM-based psychotherapy simulations were technically feasible, demonstrated experimental fidelity, and showed theoretically concordant effects beyond directly manipulated constructs and simulation participants. These findings provide proof of concept for *in silico* psychotherapy research as a promising complementary pre-clinical experimental paradigm for hypothesis generation and testing to support accelerated intervention development. The applicability of this approach across patient populations, therapeutic modalities and model architectures requires further evaluation.

**Trial Pre-registration:** ClinicalTrials.gov NCT06813066; https://clinicaltrials.gov/study/NCT06813066?term=NCT06813066

## Introduction

Psychotherapy is a core component of evidence-based care for a broad range of mental disorders and is recommended as a first-line intervention in many clinical guidelines. Psychotherapeutic approaches have improved symptoms and functioning across conditions, ranging from depression and anxiety to more complex conditions like personality disorders, and can contribute to sustained improvements in quality of life.^1,2^ However, despite substantial advances in both psychotherapeutic and pharmacological treatments, population-level outcomes for common mental disorders appear to have stagnated. Beyond widely recognised access-related challenges, umbrella reviews and meta-analytic evaluations also suggest that average intervention effects remain modest (psychotherapies: SMD = 0.34, 95% CI 0.26–0.42; pharmacotherapies: SMD = 0.36, 95% CI 0.32–0.41, compared with placebo or treatment as usual) and that incremental gains in efficacy have been limited over recent decades, consistent with a potential ceiling in current approaches.^3^ This plateau has sharpened interest in methodological innovation that can accelerate discovery, increase experimental control, and improve mechanistic understanding in mental health intervention research.

One promising route is to expand the toolkit of health research beyond *in vivo* and *in vitro* studies to include *in silico* strategies – computational simulations that enable controlled, rapid and ethically tractable hypothesis testing. In biomedicine, *in silico* clinical trials and related modelling approaches are increasingly used to explore mechanisms, optimise interventions and reduce reliance on early-stage animal testing.^4,5^ These approaches draw on established principles of scientific simulation, including verification and validation to build model credibility and the broader practice of modelling complex systems.^6,7^ More broadly, computational attempts to emulate elements of human cognition and decision-making have a long history, ranging from simple rule-based models to Distributed Artificial Intelligence (DAI) and Multi-Agent Systems (MAS) frameworks demonstrating emergent behaviours resulting from the coordinated interplay of autonomous agents with distinct roles.^8–10^ Concurrently, advances in generative artificial intelligence (genAI) and large language models (LLMs) have made it feasible to simulate structured, context-sensitive dialogues at scale, relying on large-scale associative/stochastic inference instead of explicitly defined, sophisticated assumptions concerning cognitive or social processes.^11^ Previous studies have already proposed LLM-based patients and/or therapists as training or evaluation tools and for generating synthetic model training data. ^12–14^

Together, these developments motivate a programme we term *‘in silico* psychotherapy research’: the use of controlled, reproducible simulations of psychotherapeutic interactions to interrogate, and ultimately refine, mechanisms of therapeutic action. *In silico* simulations can generate large numbers of sessions, implement precise experimental manipulations (e.g., varying the therapist’s relational stance while holding other features constant), and standardise initial simulated patient presentations. Such control can isolate variables that are difficult to randomise, conceal or stabilise in conventional psychotherapy trials, while providing a platform to generate testable predictions and prioritise hypotheses for subsequent human research.^4^ In addition, simulated data may support methodological development, such as automated assessment of treatment fidelity and process indicators.^15,16^

At the same time, the use of LLMs in health-related research raises important concerns about validity, equity and safety. Recent commentaries and evaluation frameworks emphasise that LLM outputs must be prospectively validated against expert benchmarks, and that evaluation workflows should be transparent, reproducible and resistant to bias.^16–19^ Empirical studies further indicate that LLM outputs may vary systematically with sociodemographic descriptors even when clinical content is held constant, highlighting potential fairness risks.^20,21^ Together, these observations argue for study designs that (a) constrain LLM tasks to well-specified therapeutic principles; (b) include manipulation checks to ensure the model is behaving as intended (for example adhering to a particular therapeutic style); (c) benchmark LLM-based evaluations against human ratings; and (d) whenever possible, anchor simulations to real-world clinical materials.^16^

Motivational interviewing (MI) offers a particularly suitable starting point for this research programme, and has been used in previous LLM-based frameworks targeting model development and quality assessment.^12,13^ MI is a structured, widely disseminated approach with evidence of efficacy across settings and target behaviours, including mental health, and a broad range of medical and primary-care contexts.^22–24^ Importantly, MI is supported by an established fidelity framework, the Motivational Interviewing Treatment Integrity code 4.2 (MITI), which operationalises therapist behaviours and global ratings of therapist style, thereby enabling standardised evaluation of MI-consistent practice.^25^

Beyond specific techniques, psychotherapy research consistently underscores the contribution of common factors, including relational and interpersonal qualities of therapists that cut across theoretical orientations.^26^ Meta-analytic syntheses indicate that alliance, empathy, positive regard or affirmation and related constructs are robustly associated with treatment outcomes and are plausible contributors to therapeutic change.^27–29^ These constructs are also amenable to operationalisation via observer-rated scales, making them suitable targets for controlled experimental manipulation and for linking process variation to outcome-relevant indicators in both conventional and simulated designs.

In the present study (pre-registered at ClinicalTrials.gov, NCT06813066), we sought to establish an initial proof-of-concept for an “*in silico* psychotherapy research” paradigm, in which a simulated therapist conducts MI with a simulated patient across standardised case vignettes and therapist personas. The implemented approach orchestrates LLM-based role-corresponding components and relies primarily on the associations implicitly captured in the model weights, intentionally foregoing explicit cognitive architectures or complex social modelling to prioritise experimental scalability and tractability over internal cognitive fidelity. We thereby pursued two main objectives. First, we assessed feasibility by evaluating whether LLM-based simulations are able to reliably generate coherent, role-consistent and clinically plausible psychotherapeutic dialogues that adhere to core MI principles, operationalised primarily through observer-rated MI treatment integrity, the frequency of severe simulation failures (e.g., looping, role deviation, incomprehensibility) and the perceived credibility of the participants. Second, we assessed experimental sensitivity by manipulating the simulated therapist’s common factors (high versus low empathy/warmth, affirmation/positive regard and alliance-building behaviours) while holding all other instructions constant, and testing whether this manipulation produced systematic differences in MI process indicators, in proxy outcomes of simulated patient change, or in linguistic characteristics. To contextualise the ecological plausibility of the simulations and to benchmark the evaluation workflow, we additionally compared simulated sessions to published real-world MI transcripts with matching demographic and clinical patient characteristics, and examined agreement between LLM-based ratings and blinded human assessments in a stratified random subsample.^13^

If further validated, this paradigm could provide a scalable preclinical platform for hypothesis generation and method development before resource-intensive human stud_ies._4,17,19–21

## Methods

### Study design and preregistration

Tis interventional study was conducted as a proof-of-concept for the proposed *in silico* psychotherapy research paradigm and was pre-registered under ClinicalTrials.gov (NCT06813066; https://clinicaltrials.gov/study/NCT06813066?term=NCT06813066; 2025-02-06) to facilitate transparency, prospective specification, discoverability, reproducibility, and comparison with human studies. It evaluated whether LLMs can be used to simulate complete therapy sessions adhering to specific methodological guidelines and sensitive to theoretically and/or clinically relevant experimental manipulations in a controlled, reproducible simulation framework.

As a test case, we manipulated the level of therapist-exhibited common therapeutic factors (CTF) in LLM-simulated Motivational Interviewing (MI) sessions. The simulations followed a balanced fully crossed factorial design with a binary experimental condition, 8 therapist and 8 patient personas and four replications for each possible combination (see Figure 1 and Supplementary Table ST3). The set of eight publicly available transcripts of real-world MI sessions underlying the eight patient vignettes served as matched comparators.

**Figure 1.**
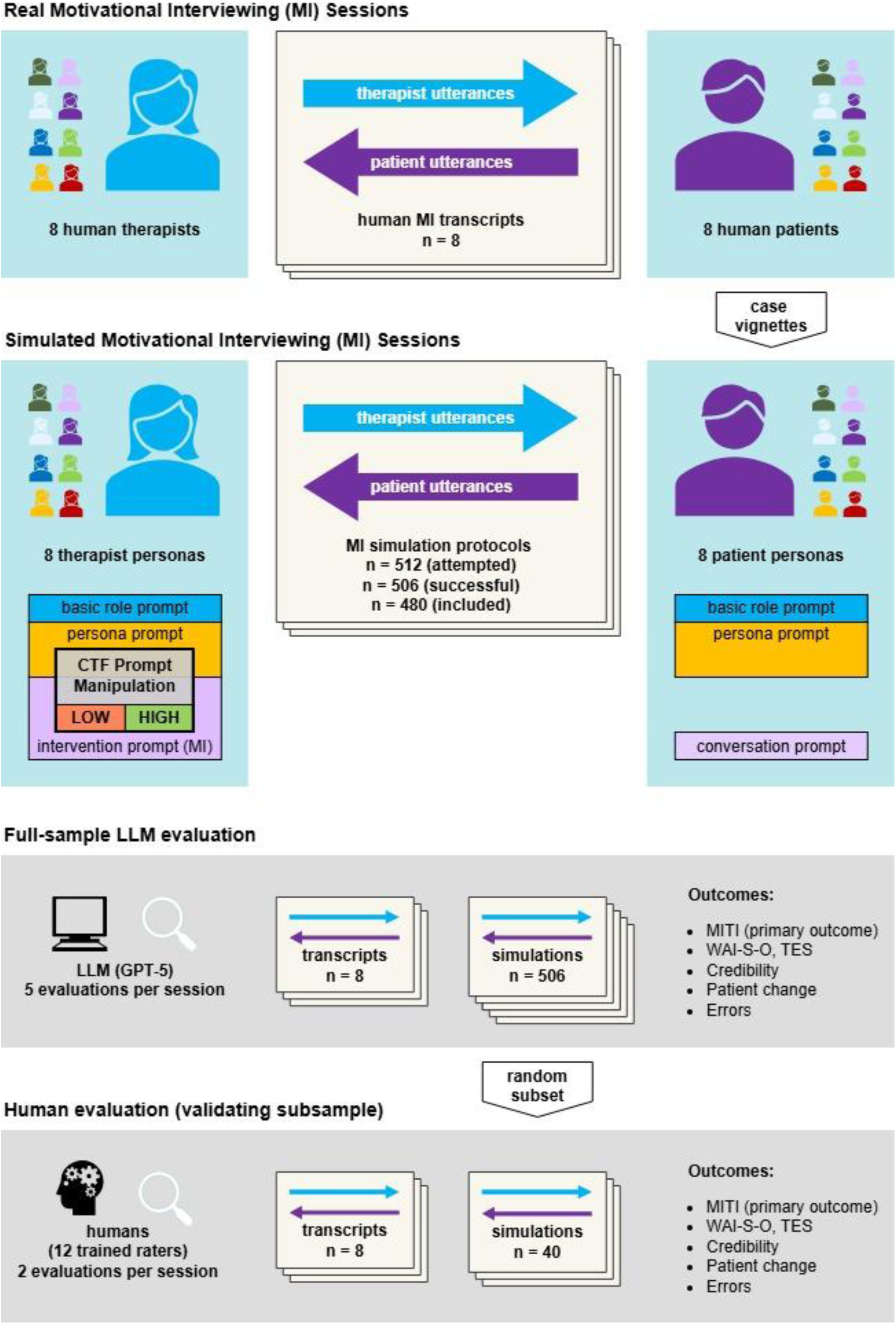
Study design and evaluation workflow for in silico motivational interviewing simulations Notes: We simulated motivational interviewing (MI) sessions using a multi large language model (LLM) architecture in which an LLM-based therapist and an LLM-based patient interacted based on predefined therapist personas and patient vignettes. Simulations were generated under two experimental conditions that manipulated the therapist’s common factors (high versus low). All simulated sessions and a set of publicly available real-world MI transcripts were evaluated using GPT-5 with repeated ratings per transcript and prespecified scoring instructions. A stratified subset of simulated sessions plus the real transcripts underwent blinded human rating to benchmark LLM-based evaluation. Abbreviations: AI, artificial intelligence; LLM, large language model; MI, motivational interviewing; MITI, Motivational Interviewing Treatment Integrity; TES, Therapist Empathy Scale; WAI-S-O, Working Alliance Inventory–Short Observer form, CTF, common therapeutic factors.

All simulations were launched and executed without human interaction after batch initiation, following a programmatically randomised serial order.

### Ethics statement

The need for an ethical review and approval was waived by the Commission for Research Ethics and Research Security, Trier University, Germany (EK Nr. 51/2024) for the entire study. Informed consent was obtained from the human raters prior to the evaluations and was not required elsewhere as this study was based on publicly available and synthetic data, and did not involve interaction with, or the collection of new data from patients.

### Simulation framework and implementation

Simulations were scripted in python and were batch-executed locally in a secure high-performance computing (HPC) environment (sciCORE+, University of Basel) relying on an open-weight conversational LLM (Llama-3.3-70B-Instruct, 8-bit quantisation). Implementation details not reported here are described in the Supplementary Methods SM1.

#### LLM-based simulation architecture and dialogue generation

Each simulated session was generated by linking multiple LLM-based agents with distinct roles. Distinct agents generated therapist utterances, patient utterances, and brief therapist backchannels, respectively; a dedicated agent monitored each exchange for session closure to trigger simulation termination. All LLM-based agents were instantiated from the same base model; role differentiation was achieved entirely through system and role prompts. Generation parameters were prespecified and held constant within each LLM-based simulation element across all sessions. Sessions were recorded in aligned patient-only, therapist-only and full transcript versions. Supplementary Figure F1 explains the schematic pipeline.

#### Prompting

Prompts were structured to (i) establish role identity (patient vs therapist), (ii) provide persona- and vignette-specific context, (iii) specify dialogue style constraints compatible with psychotherapy transcripts, and in the LLM-based therapist, (iv) to operationalise MI-consistent practice and (v) to display high or low levels of CTF depending on the experimental manipulation.

#### Patient and therapist personas

Eight standardised vignettes for the patient personas were constructed based on eight publicly available MI transcripts (see Supplementary Table ST1). Vignette texts were used exclusively as prompts to initialise the LLM-based patient and were held constant across conditions and therapist personas.

Further, eight therapist personas were developed to introduce controlled heterogeneity in non-manipulated therapist characteristics (see Supplementary Table ST2). Personas were orthogonal to the experimental manipulation: the CTF manipulation was implemented by condition-specific instruction modifications added to the LLM-based therapist prompts, whereas all other persona and role instructions were identical across conditions.

#### Quality Control during and after simulation

To reduce artefacts that commonly arise in LLM dialogue generation during simulation, each candidate utterance underwent a rule-based validity check prior to being appended to the transcript (see Supplementary Figure SF1). Following completion, the automated LLM evaluation procedure repeatedly screened transcripts for the following severe simulation errors: abrupt termination without closure, looping or repetitive patterns, major role deviation (e.g., patient adopting therapist role), sustained language switching away from English or incomprehensibility, or any other severe error. Protocols flagged with any of the above errors in any of the screening rounds were manually inspected. Simulations containing confirmed errors were excluded from downstream analyses.

### Interventions and comparators

MI was selected as the proof-of-concept intervention for this simulation study due to its clear structure, broad empirical background, wide area of applications and existing evaluation framework.^22–24^ In this simulation study, the MI intervention was administered to LLM-based patients through LLM-based therapists. The LLM-based therapists were provided with detailed instructions about the MI intervention in their system prompts.

The MI intervention was delivered in two versions depending on the experimental condition, with LLM-based therapists instructed to display different levels of CTF:

1. *High levels of CTF*: In this condition, the LLM-based therapist was prompted to exhibit high levels of empathy, warmth, affirmation or positive regard, and the capacity and readiness to build rapport. This configuration was intended to foster a therapeutic environment that enhances patient engagement and trust. High CTFs are associated with effective psychotherapy, as they contribute to a stronger therapeutic alliance and better patient outcomes.^27,30^
2. *Low Levels of CTF*: In this condition, the LLM-based therapist was prompted to exhibit low levels of CTF. This approach aimed to explore the impact of a less supportive and empathetic therapist behaviour on the dynamics of psychotherapy sessions. In real-world scenarios, lower levels of these behaviours can lead to reduced patient engagement and a weaker psychotherapeutic alliance, potentially hindering the therapeutic process.

### Outcomes

#### Primary outcome

The primary outcome was the quality of the simulated intervention and the simulation itself, operationalised as MI treatment integrity and assessed with the MITI 4.2 global ratings including cultivating change talk, softening sustain talk, partnership, and empathy. ^25^

#### Secondary outcomes

Secondary outcomes were defined a priori as follows:

1. Severe simulation errors: prevalence of non-technical errors meeting exclusion criteria (see Quality Control) and technical failures (e.g., timeouts) during generation.
2. Manipulation checks: therapist empathy and working alliance assessed using an observer-rated Therapist Empathy Scale (TES; adapted by excluding items requiring acoustic information, see Supplementary Materials) and the Working Alliance Inventory – Short Observer form (WAI-S-O).^31,32,33^
3. Observer-inferred proxy patient improvement: change in perceived importance and confidence for the target behaviour, operationalised using an observer-rated adaptation of the MI “importance” and “confidence” rulers, based on patient utterances alone, scored pre-session and post-session on a scale from 1 (not important at all) to 10 (very important). Change scores were computed as post minus pre values.^34^
4. Credibility of simulated behaviour: perceived authenticity of therapist and patient behaviour rated on a 0–10 scale (0 = certainly simulated; 10 = certainly human/real; raw LLM-rated scores were inverted for technical reasons).^35^
5. Linguistic metrics: quantitative and qualitative text properties for therapist and patient utterances, including counts of turns/utterances, sentences, words, syllables, characters and derived indices of lexical diversity (measure of textual lexical diversity; MTLD).^36^
6. Turn-takings: number of utterances/exchanges between therapist and patient within each session, derived from full dialogue protocols.

### Evaluation procedures

To ensure efficient, robust and benchmarked evaluation, all outcomes were assessed using two complementary approaches: a scripted independent LLM-based rating procedure applied to all sessions, and blinded human ratings on a stratified subsample, the latter serving to validate LLM-based scores against independent expert judgement.

#### LLM-based ratings

All primary and secondary outcomes requiring judgement (MITI 4.2 global ratings, patient change rulers, credibility ratings, TES, WAI-S-O) were scored using GPT-5 as an evaluation model via independent scripted API calls. Importance and confidence scores were evaluated based on patient utterances only. To improve robustness, each transcript was rated five times per outcome. Scores were retrieved automatically from the output and underwent formal validity control steps; missing or erroneous scores triggered up to five repeated evaluation attempts. For implementation details see Supplementary Methods SM1.

#### Human rater validation

To benchmark the LLM-based evaluation procedure against expert assessment,^17^ we randomly selected a stratified subsample of 40 simulated protocols (representing 7.9% of all LLM-based protocols) to ensure balance across CTF conditions and patient vignettes. The eight real-world MI transcripts were added to this set, yielding a total validation corpus of 48 elements. Twelve human raters independently assessed assigned sessions in a randomised order, each session was evaluated by two raters. Human ratings followed the same operational definitions and scoring instruments used for LLM-based evaluation (MITI 4.2 global ratings, TES, WAI-S-O, patient change rulers, and credibility ratings), using standardised, detailed instructions in a fixed sequential order. Change ruler scores were evaluated first, based on a transcript version containing patient utterances only; all subsequent outcomes were evaluated based on the full transcript.

To ensure sufficient competence, human raters were required to hold a university degree in psychology encompassing standardised clinical and counselling competencies in accordance with current German licensing regulations (PsychThApprO). Further, they were provided with a written detailed description of the goals, principles, and stages of Motivational Interviewing (MI) prior to the rating tasks. They were recruited via the university’s digital black board (Uni-Verteiler) and received €100 compensation for their participation.

#### Text analytics

Text-based outcomes were computed using a rule-based pipeline in R, including to-kenisation and aggregation of word counts and related indices. Lexical diversity was quantified using MTLD. Text metrics were computed separately for therapist and patient as well as at session level.

### Sample Size

Given the proof-of-concept *in silico* framework, no directly comparable effect size estimates were available to inform an a priori power calculation. We therefore used a precision-based sample-size criterion and selected a balanced design of 512 simulated sessions (8 patient vignettes × 8 therapist personas × 2 CTF conditions × 4 replicates). In simulations allowing for up to 20% attrition, this design yielded an expected 95% CI half-width of approximately 0.195 SD units for the condition effect, not surpassing a small effect size of interest (d=0.2).

### Randomisation

We prespecified a fully crossed design (8 patient vignettes × 8 therapist personas × 2 CTF conditions × 4 replicate sessions per pairing). Simulation runs were executed in a programmatically randomised order to mitigate potential serial or resource-related effects. For the human-validation corpus, we used block randomisation with documented timestamp-based seeds for stratified subsample selection, rater assignment, and ordering. This approach maximised comparability between the full sample and the subset and ensured that each rater received a balanced mix of protocols across different conditions and vignettes, enhancing the integrity and validity of our comparative analysis.

### Blinding

Given the nature of this simulation study, traditional concepts of blinding (masking) do not apply as typically understood in human trials. Nevertheless, human evaluators were blinded both to the condition (low vs. high CTF) and to the simulated or real nature of the sessions. Similarly, the evaluation LLM (GPT-5) was informed neither to the CTF condition nor to the simulated or real nature of the sessions. As LLM rating tasks were independent, the model was not informed about other transcripts, outcomes or prior ratings. Further, the therapist and patient system prompts did not contain information that a therapist or patient would not usually have prior to a real psychotherapy session. Neither the LLM-based therapist nor the LLM-based patient was informed about the experimental setting. Importantly, LLM-based patients were initiated identically across conditions and therapist personas and were not informed about any specific features of the simulated therapist.

### Statistics

We conducted all statistical analyses in R (R Foundation for Statistical Computing, Vienna, Austria, see Supplement for packages). The core analytic framework was determined before inferential analysis; model-specification refinements and post-hoc robustness analyses are described in the supplement. For simulation-based analyses, the unit of analysis was the simulated session transcript. We summarised feasibility as counts and proportions of technical failures (e.g., timeouts) and severe simulation errors identified during post hoc screening. We excluded transcripts flagged with severe simulation errors from downstream inferential analyses and performed complete-case analyses for each endpoint.

For outcomes scored by the evaluating LLM and/or human raters (MITI 4.2 global ratings, TES, WAI-S-O, credibility ratings, and importance/confidence rulers), we analysed transcript-level aggregated mean scores. We reported descriptive statistics by CTF condition (high vs low) as mean (SD) and, where informative, median (IQR). We based distributional assessments on visual inspection (histograms and Q–Q plots); we treated formal normality tests as descriptive given their sensitivity in large samples. All confidence intervals (CIs) were 95% and all tests were two-sided.

#### Primary outcome (MITI 4.2 global ratings)

We estimated the effect of CTF condition on each MITI 4.2 global rating using linear mixed-effects models reflecting the crossed design. Each model included CTF condition as a fixed effect and random intercepts for patient vignette and therapist persona:

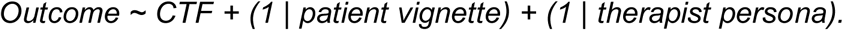

We additionally tested allowing the CTF effect to vary across patient vignettes via a random slope. We used lmerTest for fixed-effect inference (Satterthwaite degrees of freedom) and reported adjusted mean differences for CTF (high vs. low) with 95% CIs and p values. When residual diagnostics suggested material departures from homoscedasticity or approximate normality, we examined robustness using transformation-based approaches and/or alternative estimators as described in the supplement.

#### Secondary outcomes

We analysed manipulation checks (TES and WAI-S-O total and subscales), credibility ratings (0–10; higher values indicating higher perceived credibility/human-likeness in the analyses), proxy patient change (post–pre change in importance and confidence rulers) and text-derived metrics (e. g. total number of utterances and words, mean word count per utterance, and MTLD lexical diversity computed separately for therapist and patient text) using the same mixed-effects framework and contrast definition (high minus low CTF).

#### Contextual comparison with published real-world MI transcripts

To contextualise ecological plausibility, we compared simulated sessions with a set of published MI transcripts at the vignette level. For each vignette, we aggregated simulated outcomes across sessions corresponding to that vignette and paired the vignette-level mean value with the corresponding published transcript. We planned paired t-tests when paired differences were approximately symmetric and non-parametric tests otherwise. We report paired differences with bootstrapping-based 95% CIs (5000 resampling), as well as standardised paired effect sizes (Cohen’s *d*) and permutation-based p-values for comparability.

#### Reliability and validation

We quantified reliability of repeated LLM ratings using intraclass correlation coefficients (ICCs), treating repeated ratings as independent “raters” and reporting ICCs for for the mean of repeated ratings (absolute agreement forms as appropriate). In the human-validation corpus, we assessed agreement between LLM evaluations and human ratings using ICCs for continuous outcomes and complementary indices (weighted kappa and/or Krippendorff’s alpha where an ordinal interpretation was meaningful). We examined systematic bias and limits of agreement using Bland–Altman analyses and, where feasible, reported attenuation-corrected agreement estimates. Further, we estimated CTF effects on all LLM-evaluated outcomes in based on human ratings.

#### Post-hoc robustness analyses

To assess attrition bias from post-simulation exclusion, we re-run the primary mixed-effects models including all successfully saved simulations. Additionally, to inspect whether CTF effects were detectable beyond quantitative conversation structure differences, we repeated the same analyses controlling for length characteristics (mean patient and therapist utterance length and total utterance count) as fixed effect covariates. Further, for quantitative text metrics, we estimated multiplicative effects using mixed effect models with a log scaling and an otherwise identical fixed and random effects structure.

### Harms

The simulated interventions did not involve human participants or newly collected human data, and all real MI transcripts were collected from publicly available sources. Accordingly, direct intervention-related adverse events or harms were not applicable. Careful interpretation of the simulation-based findings is nevertheless important to prevent downstream harms from unsubstantiated clinical or resource allocation decisions.

## Results

Across the fully crossed design, 512 *in silico* motivational interviewing (MI) sessions were initiated (Figure 2 and Supplementary Table ST3). Six simulation runs (1.17%) failed for technical reasons (timeouts), all in the low CTF condition. Of the 506 successfully generated session protocols, 26 (5.14%) were excluded post hoc due to severe non-technical simulation errors (e.g., looping/loss of progression or abrupt termination without plausible closure), yielding a final analysis set of 480 simulated sessions (237 low CTF; 243 high CTF). The distribution of CTF condition, patient vignette, and therapist persona levels across the attempted simulations remained near-balanced among successful and included simulations (see Supplementary Table ST3).

**Figure 2.**
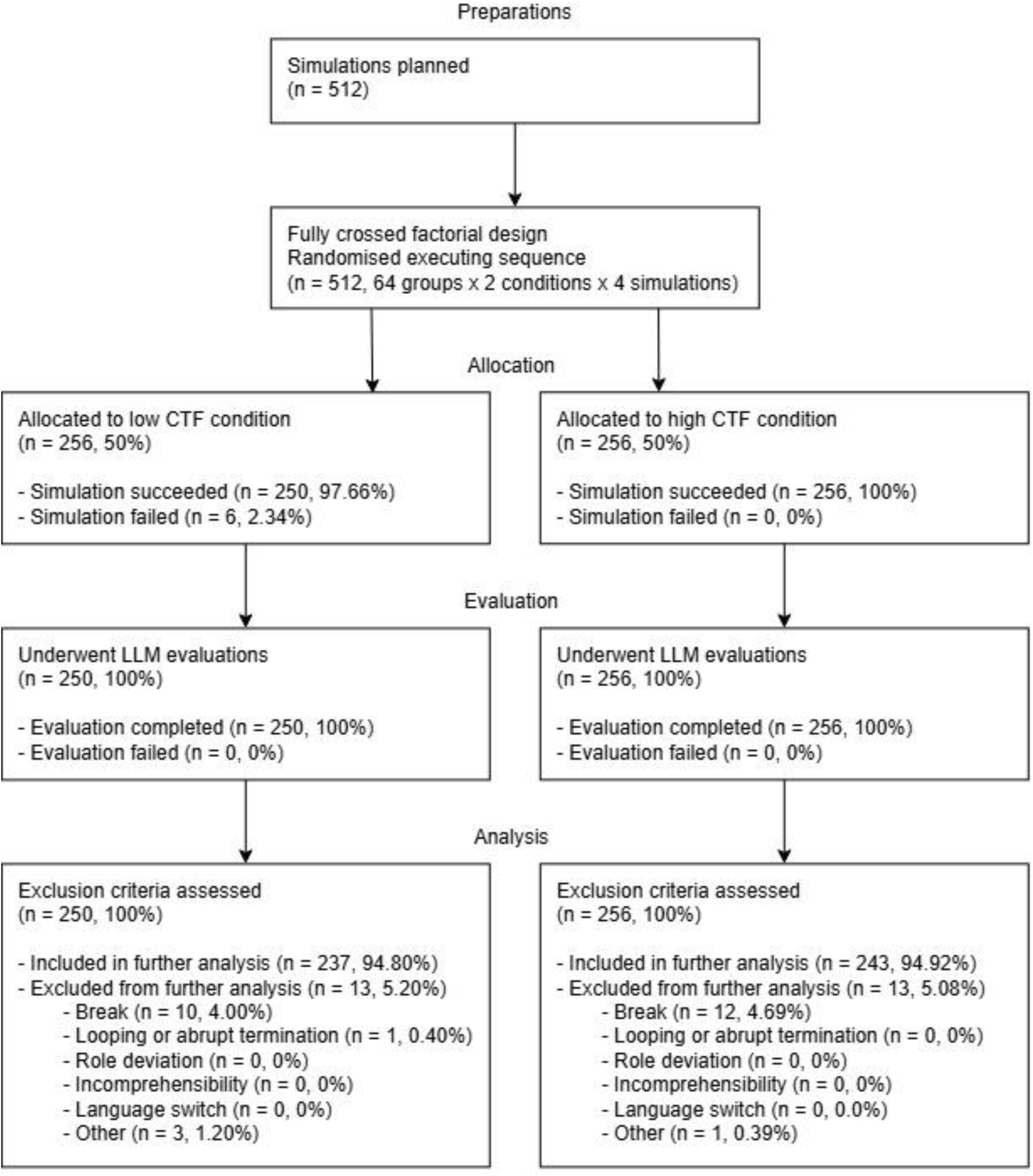
CONSORT-style flow diagram of *in silico* session generation, exclusions, and analysis sets. *Notes: The figure summarises the number of attempted simulations, successful generations, technical failures, post hoc exclusions due to severe simulation errors, and the final analysis sample by experimental condition (high versus low common therapeutic factors, CTF). Severe simulation errors comprised looping or loss of conversational progression, major role deviation, sustained language switching away from English, incomprehensibility, or abrupt termination without plausible closure. Abbreviations: CONSORT, Consolidated Standards of Reporting Trials; CTF, common therapeutic factors; LLM, large language model*.

### Descriptive outcomes by CTF condition

Descriptive statistics for primary and secondary outcomes in the included simulated sessions are summarised in Supplementary Table ST7, and corresponding values for the real MI transcripts are shown in Table 1. Overall, simulated sessions exhibited clear sensitivity to the experimental manipulation on MI process integrity (MITI 4.2 global ratings, see Figure 3A) and on manipulation-check measures (therapist empathy and working alliance, see Figure 3B), while showing smaller differences on proxy simulated patient change and credibility outcomes.

**Figure 3.**
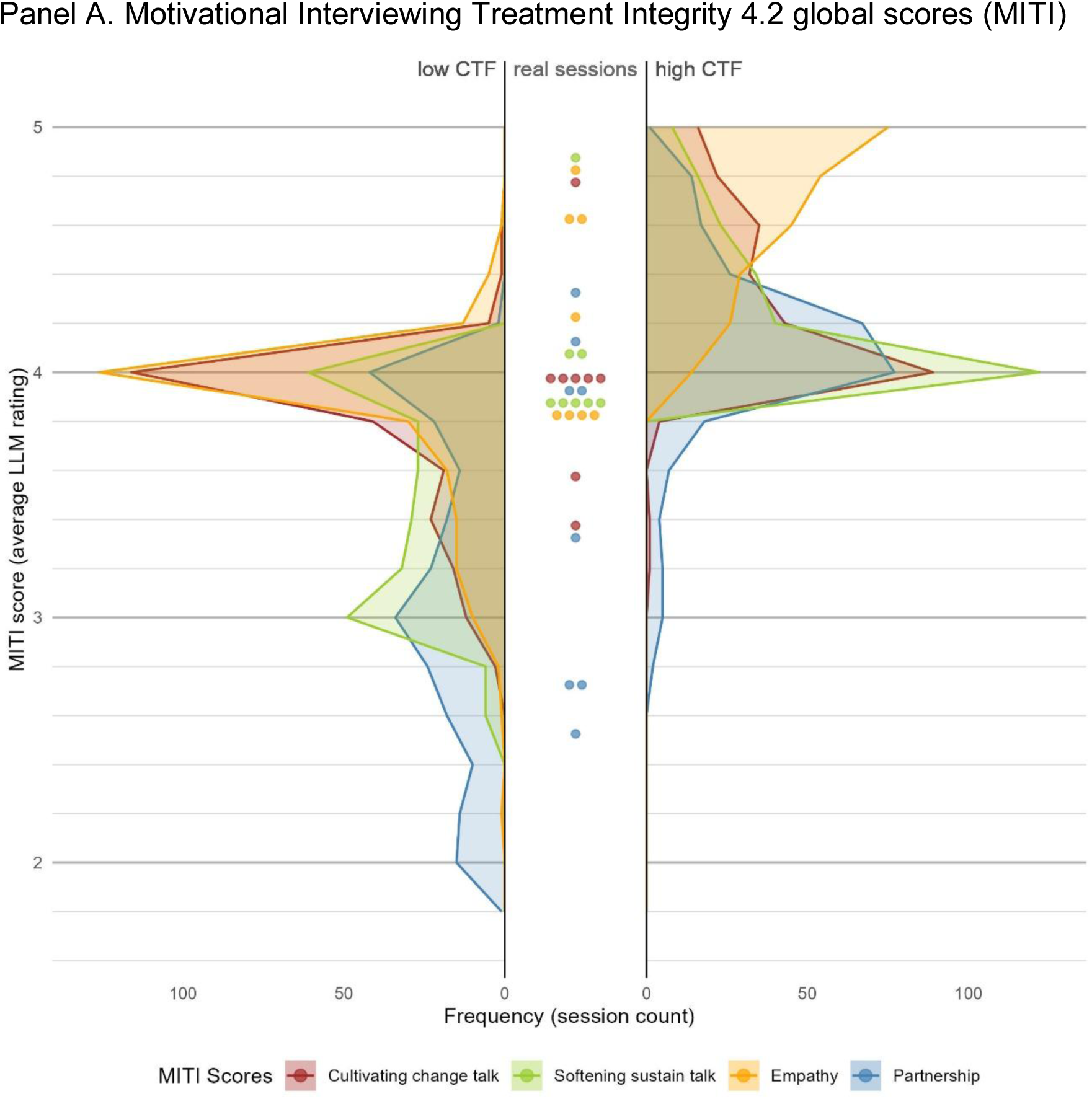
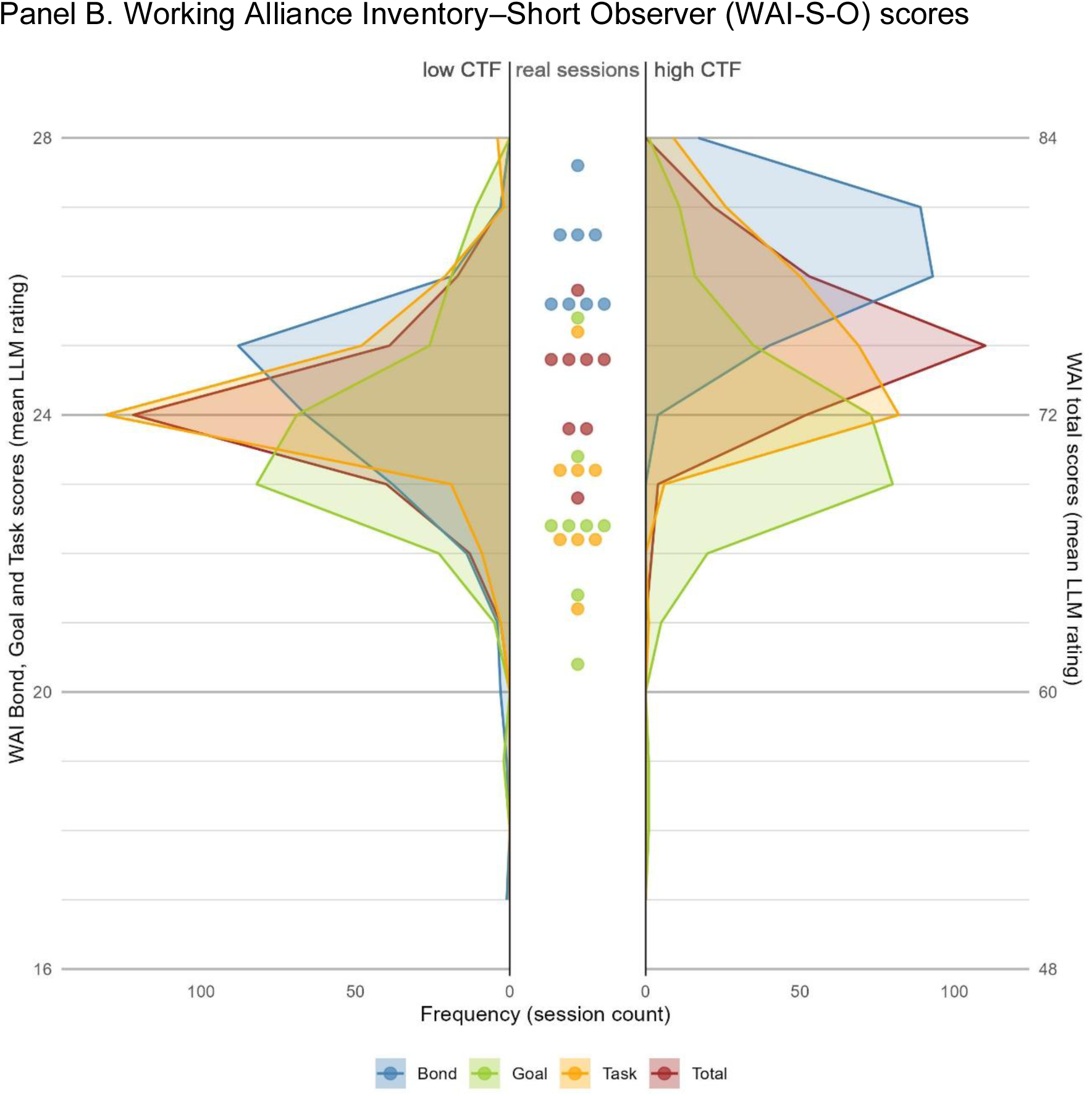
Distribution of Motivational Interviewing Treatment Integrity and Working Alliance Inventory scores in simulated sessions and real transcripts. *Notes: Panel A shows distributions of GPT-5-rated MITI 4.2 global scores for simulated sessions generated under low (n = 237) and high (n = 243) common factors conditions, alongside scores from published real-world MI transcripts (n = 8). Panel B shows corresponding distributions for the working alliance rated using the WAI-S-O (total and subscales: goal, task, bond). Plotted score values reflect transcript-level mean values aggregated across repeated GPT-5 ratings. “Real sessions” refers to the set of published MI transcripts rated using the same evaluation prompts, where each dot represents one transcript. For readability reasons, WAI-S-O scores were rounded, and real session score stacks were visually dodged around the actual value. Abbreviations: CTF, common therapeutic factors; LLM, large language model; MI, motivational interviewing; MITI, Motivational Interviewing Treatment Integrity; WAI-S-O, Working Alliance Inventory–Short Observer form*.

**Table 1.**
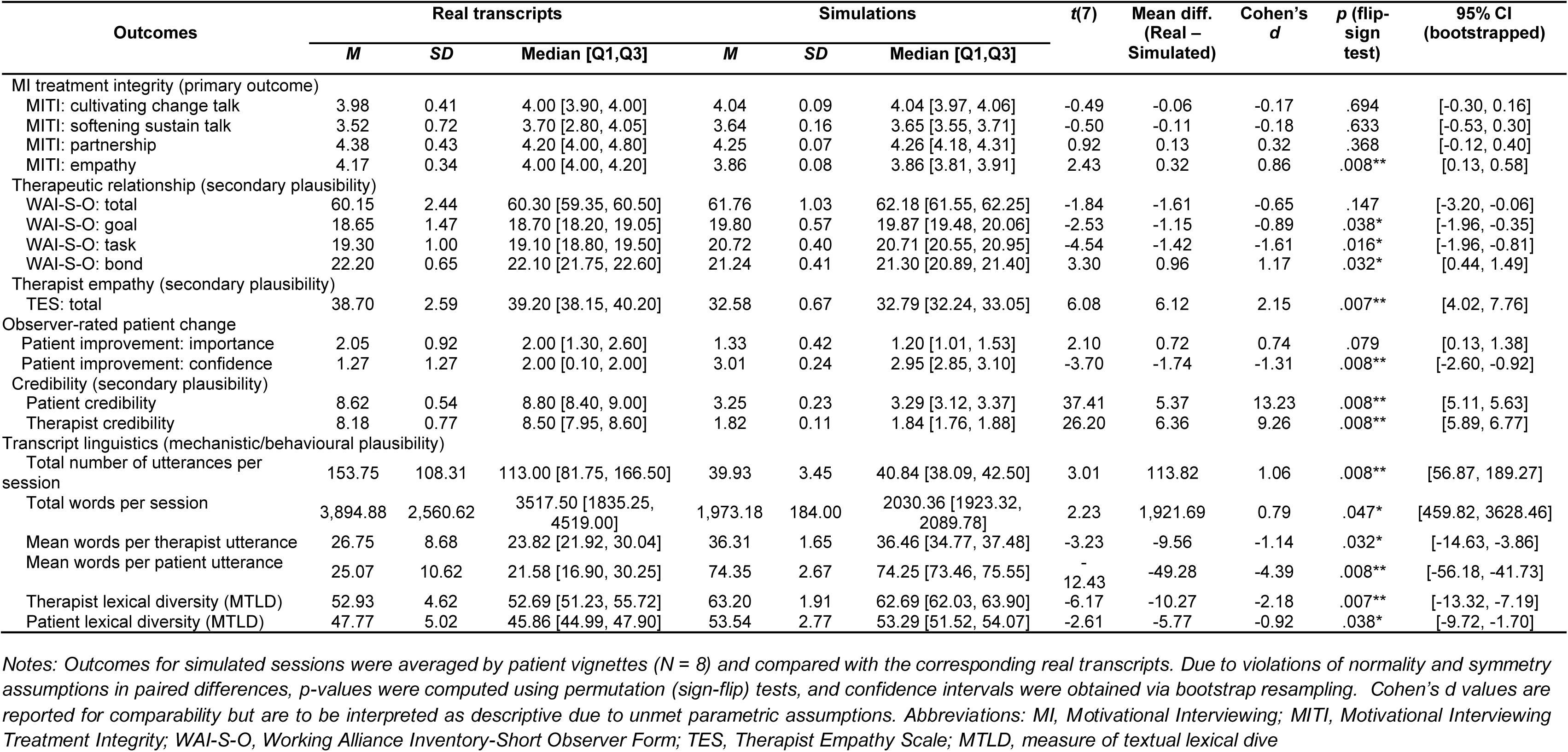
Differences in outcome variables between simulated and real sessions.

### MI treatment integrity (MITI 4.2 global ratings)

The CTF manipulation produced consistent and substantial differences in MI treatment integrity on all four MITI global domains, including the MI-specific scales “Cultivating change talk” and “Softening sustain talk”. Linear mixed-effects models with random intercepts for patient vignette and therapist persona showed adjusted mean differences (high minus low CTF) ranging from 0.54 to 0.95 points on the 1–5 (β) scale (Figure 3, Panel A and Figure 5, Panel A; full model parameters in Supplementary Table ST4).

### Manipulation checks: therapist empathy (TES) and working alliance (WAI-S-O)

Manipulation checks confirmed that the intended relational stance differences were strongly expressed in the simulated therapists’ behaviour (Figure 5, Panel B; Supplementary Table ST4). High CTF sessions showed markedly higher TES scores than low CTF sessions. High CTF sessions also scored higher on overall working alliance (WAI-S-O), fuelled by differences in the task and bond, but not in the goal sub-scores.

### Proxy simulated patient change: importance and confidence rulers

Observer-rated proxy change (based on simulated patient utterances within the transcript) showed selective sensitivity to the therapist manipulation (Figure 5, Panel C; Supplementary Table ST4). The confidence ruler change was stronger in simulated patients in high CTF simulations than in low CTF simulations, while the importance ruler change did not indicate meaningful condition differences.

### Credibility of simulated therapist and patient behaviour

Perceived credibility/human-likeness (0–10 scale; higher values indicating more credible behaviour) differed modestly by condition for the simulated therapist but not for the simulated patient (Figure 4, Panel C; Supplementary Table ST4). High CTF sessions received slightly higher therapist credibility ratings than low CTF sessions, while patient credibility ratings were essentially unaffected.

**Figure 4.**
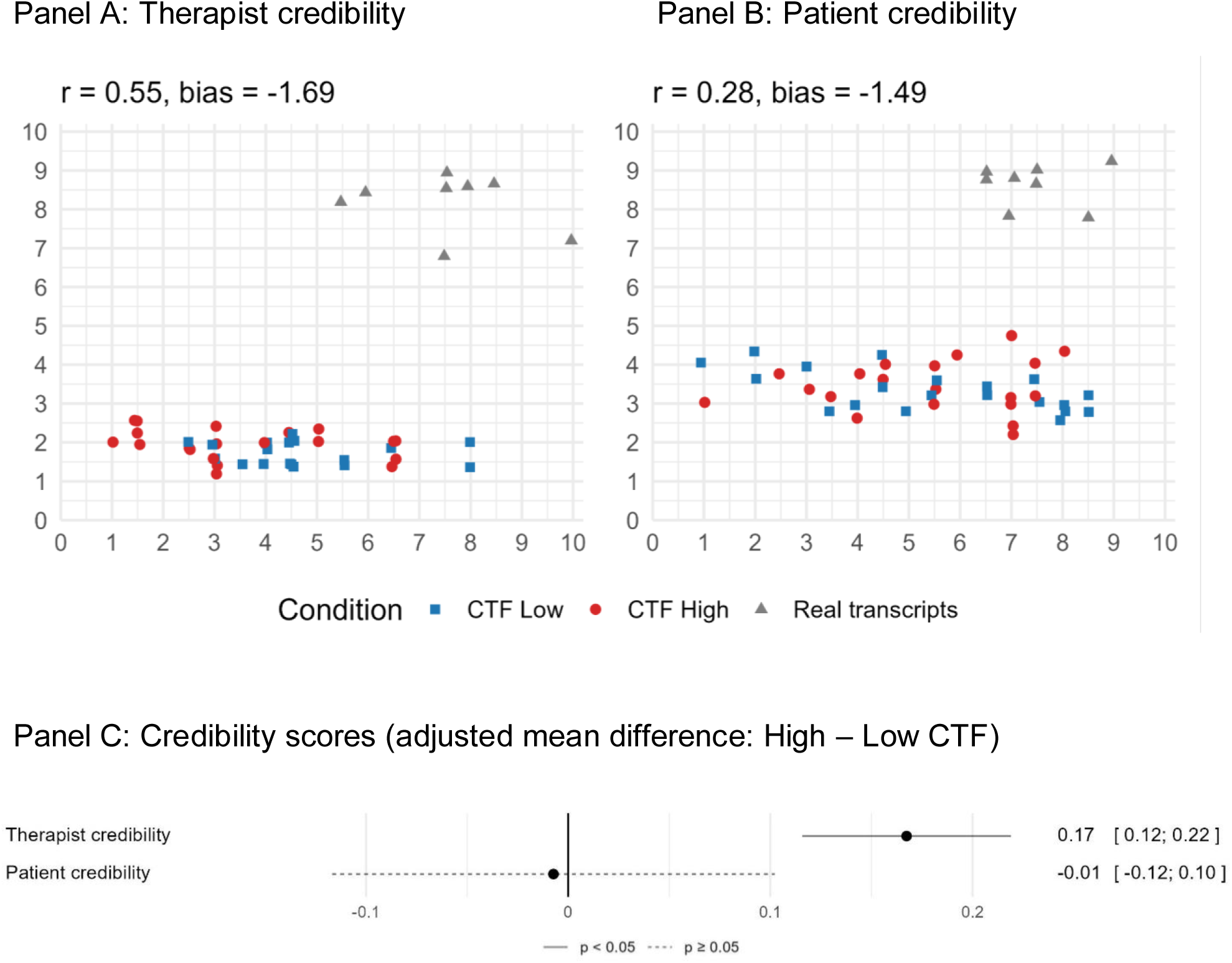
Credibility ratings: LLM versus Human Agreement and High versus Low Common Therapeutic Factors (CTF) Effects. *Notes: Credibility ratings of simulated and real sessions and agreement between LLM and human raters. Panels A and B display transcript-level credibility ratings for therapist and patient behaviour, comparing LLM ratings to blinded human ratings in the validation corpus (simulated subsample plus real transcripts). The Pearson correlation (r) and the mean rating bias (LLM minus human ratings) are provided. Panel C depicts differences between high and low common therapeutic factors (CTF) conditions in credibility ratings for all simulated sessions, LLM-evaluated, with 95% confidence intervals. Abbreviations: CI, confidence interval; CTF, common therapeutic factors; LLM, large language model*.

**Figure 5.**
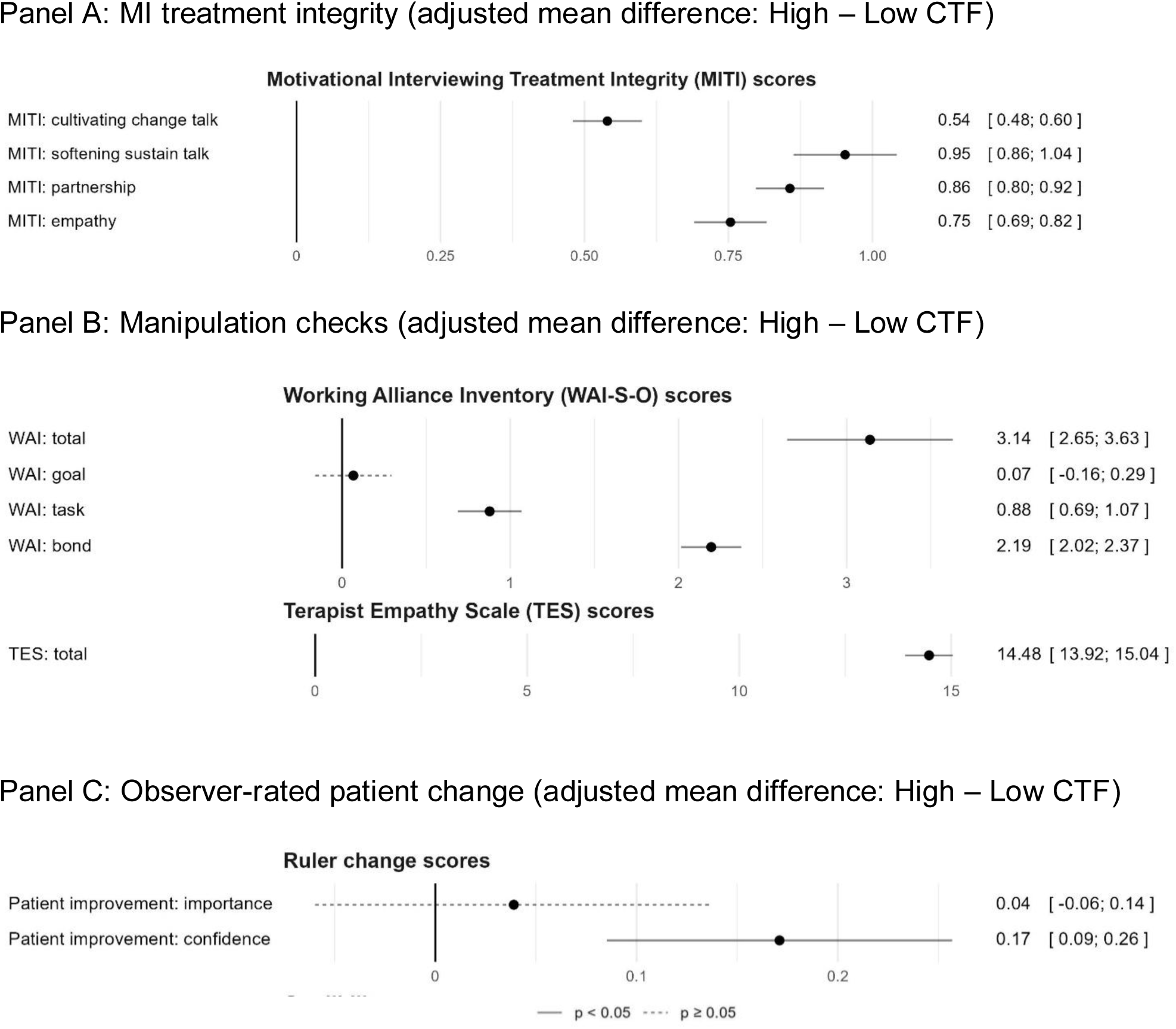
Effects of common therapeutic factors (CTF) manipulation on MI integrity, therapeutic relationship, and patient change. *Notes: Effect of the common therapeutic factors (CTF) manipulation on MI integrity, therapeutic relationship, and patient change, based on LLM evaluations. Forest plot shows adjusted mean differences (high CTF minus low CTF) with 95% confidence intervals from linear mixed-effects models accounting for clustering by patient vignette and therapist persona. Panel A reports MI treatment integrity (MITI 4.2 global ratings: cultivating change talk, softening sustain talk, partnership, empathy). Panel B reports manipulation checks (TES total score and WAI-S-O total and subscale scores). Panel C reports observer-rated proxy patient change (pre–post change in importance and confidence rulers). Abbreviations: CI, confidence interval; CTF; common therapeutic factors; MI, Motivational Interviewing; MITI, Motivational Interviewing Treatment Integrity; TES, Therapist Empathy Scale; WAI, Working Alliance Inventory; WAI-S-O, Working Alliance Inventory–Short Observer form*.

### Transcript length, turn-takings, and linguistic characteristics

The CTF manipulation also produced systematic differences in quantitative text characteristics (Figure 6; Supplementary Table ST4). Compared with low CTF sessions, high CTF sessions were overall shorter, containing markedly fewer but longer utterances, particularly on the simulated therapist but also on the simulated patient side. Lexical diversity (MTLD) increased selectively for therapist language. Sensitivity analyses using multiplicative models for all length metrics produced very similar patterns (Supplementary Figure SF2).

**Figure 6.**
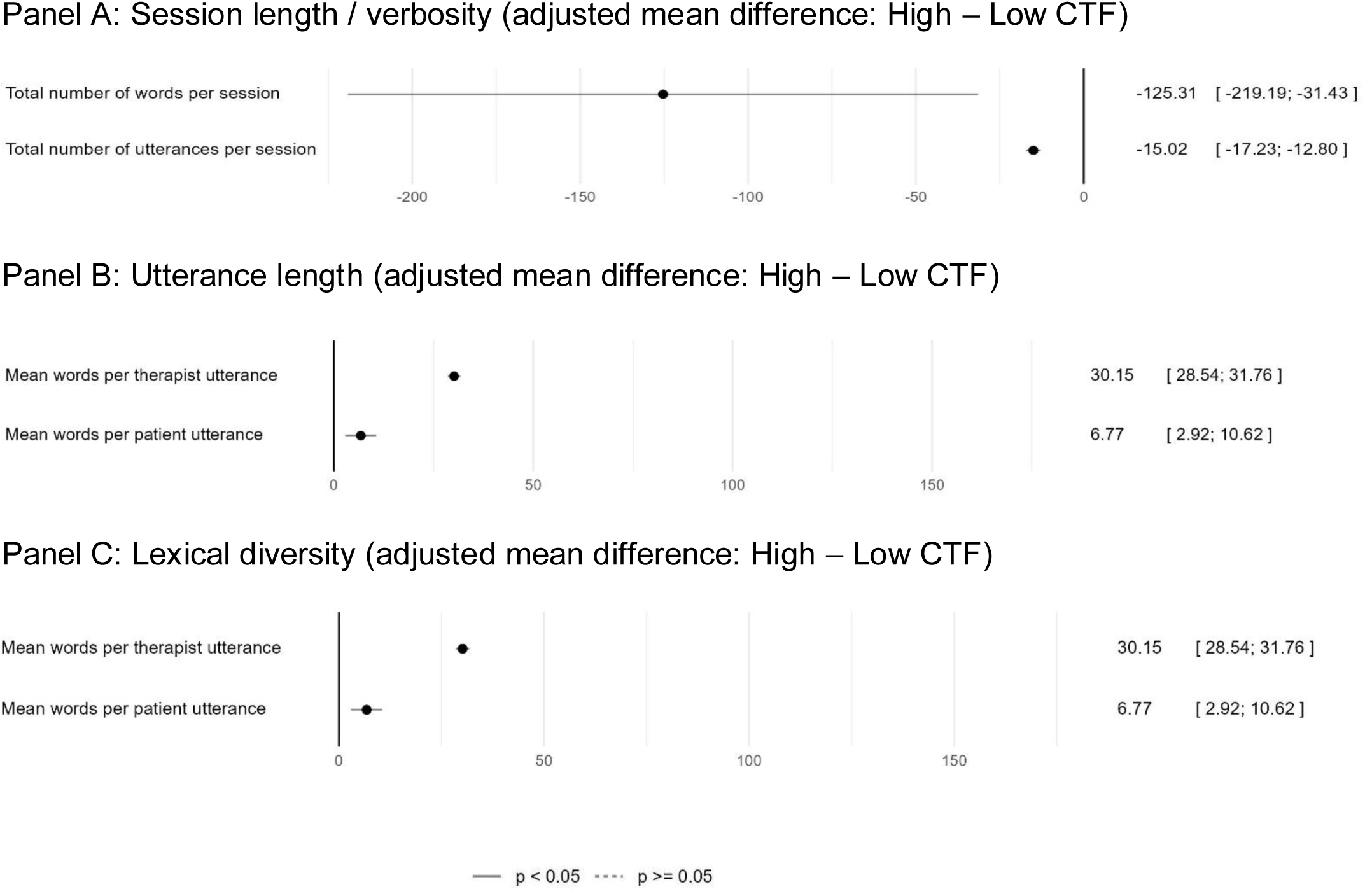
Effect of common therapeutic factors (CTF) manipulation on transcript length and linguistic characteristics. *Notes: Forest plot shows adjusted mean differences (high common factors minus low common factors) with 95% confidence intervals from linear mixed-effects models. Panel A summarises overall transcript length (total number of utterances and total word count). Panel B summarises mean utterance length (mean words per therapist utterance; mean words per patient utterance). Panel C summarises lexical diversity (MTLD) separately for therapist and patient text. Abbreviations: CI, confidence interval; CTF, common therapeutic factors; MTLD, measure of textual lexical diversity*.

### Contextual comparison with real MI transcripts

To benchmark ecological plausibility, simulated sessions were compared with eight published real-world MI transcripts (Table 1; Figures 1, 3 and 4). MITI Empathy was lower in simulations than in real transcripts whereas cultivating change talk, softening sustain talk, and partnership did not show clear differences (Table 1, Figure 3A). On relational measures, simulations showed lower TES scores than real transcripts (Table 1). For working alliance, simulations tended to show higher task and goal agreement, but lower bond compared with real transcripts (Table 1, Figure 3B), while overall differences in WAI-S-O total scores were not supported clearly.

Credibility ratings strongly differed from real transcripts (Table 1, Figure 4). Real transcripts were rated as markedly more credible than simulated sessions for both therapists and patients (Table 1), indicating detectable simulation artefacts.

Simulated sessions contained substantially fewer utterances and fewer words than real transcripts, although this finding should be interpreted cautiously given the high variability in transcript length among the eight real sessions (Table 1). Conversely, mean words per utterance were substantially higher for simulated patients and moderately higher for simulated therapists (Table 1) than for their human counterparts.

In paired comparisons, simulated patients showed numerically smaller observer-inferred importance and larger confidence changes than humans (Table 1). These findings should be interpreted cautiously because (i) the proxy change metrics are observer-inferred from transcript content, (ii) the real-transcript comparison set was small (*n* = 8), and (iii) the simulation scores were aggregated through multiple sessions, leading to more narrow distributions with fewer outliers.

### Reliability of LLM-based ratings and human validation

Repeated GPT-5 ratings (five repeats per transcript) showed high to very high consistency across all 514 rated transcripts (Supplementary Table ST5). ICCs for mean-of-five ratings ranged from 0.87 to 0.93 for MITI global ratings and most secondary judgement outcomes, with TES showing particularly high repeatability (ICC=0.99). These results indicate that, under fixed prompts and settings, the evaluation model produced stable transcript-level scores.

In the stratified human validation corpus, human raters reproduced the direction of the primary experimental contrast: high CTF sessions were rated higher than low CTF sessions on MITI global ratings and on key relational measures (Supplementary Table ST6). Agreement between LLM-based and human ratings varied substantially by outcome (Supplementary Figure SF3), with stronger concordance for TES total scores and weaker concordance for credibility ratings and some MITI talk-related constructs. Overall, these results support the use of repeated LLM ratings as a precise measurement tool within this simulation framework, while also underscoring that validity relative to human judgement is outcome-dependent and requires continued instrument-specific validation.

### Ancillary analyses: post-hoc robustness and sensitivity analyses

Repeated analysis including all successful simulations did not uncover any consequential attrition bias from post-simulation exclusion criteria. Furthermore, controlling for utterance count and length confirmed meaningful CTF effects beyond verbosity differences on MITI global ratings, manipulation check relational measures and simulated proxy patient improvement (see Supplementary Materials).

## Discussion

This preregistered proof-of-concept study demonstrates that *in silico* psychotherapy research – operationalised here as fully automated, LLM-based simulations of intervention sessions under controlled experimental manipulation – was feasible at scale and experimentally sensitive to a targeted manipulation of CTF.

Feasibility was supported by the predominance of coherent and analysable simulation transcripts with a high rate of technically successful simulations (98.83% of the 512 attempts) and a low rate of sessions excluded for severe simulation errors (5.14% of the 506 successful simulations). Together, these feasibility metrics indicate that a local, open-weight LLM deployment can support the batch generation of multi-turn psychotherapy dialogues under a standardized orchestration and quality-control workflow. Nevertheless, the observed failure modes confirm the importance of adopting verification and validation practices common in simulation science,^6,7^ implemented through runtime safeguards and explicit feasibility endpoints through this study.

The central finding is that manipulating therapist CTF produced strong, consistent changes in MI treatment integrity as measured by MITI 4.2 global ratings.^25^ In mixed-effects models accounting for clustering by patient vignette and therapist persona, high CTF sessions scored higher than low CTF sessions on all four MITI global domains. Notably, the manipulation targeted relational/interpersonal stance (empathy/warmth, affirmation/positive regard, alliance-building) yet produced sizeable shifts not only in adjacent relational MITI domains (empathy, partnership) but also in MI-specific process domains (cultivating change talk; softening sustain talk). This pattern is theoretically informative: it is consistent with the possibility that relational stance acts as an enabling condition for the enactment of other technically MI-consistent behaviours, or—within an LLM-based therapist—that prompt instructions shaping therapist “tone” and stance co-determine downstream selections of MI micro-skills and strategic responses.

The manipulated relational stance also manifested in dialogue structure differences: high CTF sessions consisted of fewer back-and-forth exchanges but markedly longer and lexically more diverse (MTLD) simulated therapist utterances.^36^ These patterns may result from a high CTF therapist language that includes more elaborated reflections, affirmations, and summarising statements that advanced the session more efficiently toward closure. This interpretation is also consistent with the adaptive termination mechanism used in the simulation pipeline: a therapist who signals consolidation and planning more clearly may trigger earlier closure decisions, thereby shortening sessions. Methodologically, this interaction between therapist style and termination logic is important: in future *in silico* studies, termination criteria should be treated as a potentially consequential design choice rather than a neutral implementation detail.

Even though simulated patients were not manipulated directly, they did exhibit selective sensitivity to therapist CTF. High CTF sessions contained produced a small but statistically supported increase in observer-inferred confidence in change (pre–post change difference: β = 0.17), whereas importance of change did not clearly differ by condition. The dissociation may reflect different sensitivity of the observation-based instruments, and/or a different reactivity to therapeutic alliance and therapist empathy.^34^

While these simulation-based results do not directly estimate clinical efficacy, they do show that a controlled manipulation of common-factor-like behaviours in an LLM-based therapist can causally influence established therapist integrity metrics and potentially relevant simulated patient change indicators in the expected direction, mirroring associations observed in traditional, in-person psychotherapy settings reported in the literature on the efficacy of MI and the role of CTF in psychotherapy.^23,27–29^ These literature-consistent condition effects in unmanipulated domains and/or agents reinforce the potential value of an *in silico* experimental platform for probing “process to response” relationships under controlled conditions, regardless of the psychological or biological fidelity of the simulations.

Benchmarking against published real-world MI transcripts provided a pragmatic check on ecological plausibility. Simulated sessions fell within broadly plausible ranges in most MI treatment integrity domains and in overall working alliance (WAI), but were below real transcripts on affective-relational measures including MITI empathy, Therapist Empathy Scale (TES) and WAI bond^25,31^, while surpassing them on more task-oriented alliance measures (WAI Task and Goal).^32^ The most salient discrepancy emerged in credibility: real transcripts were rated as markedly more human-like than simulated sessions for both therapist and patient behaviour. The dissociation also appeared in conversational structure: simulated sessions contained far fewer and longer utterances than real transcripts, especially on the patient side. These differences likely reflect known tendencies of LLMs toward verbosity and coherence-maximising monologues, which may be amplified through “transcript style” constraints discouraging back-and-forth interruptions and signals of active listening. From a translational perspective, these findings imply that while *in silico* psychotherapy simulations may already be useful for comparative and mechanistic questions under controlled manipulations, they still carry detectable artefacts and should not yet be assumed to be indistinguishable from human psychotherapy interactions.

A key methodological contribution is the combined evaluation strategy: repeated scripted LLM-based ratings to reduce stochastic evaluation noise, complemented by blinded human ratings on a stratified subsample. Repeated GPT-5 ratings showed high to very high repeatability across outcomes, indicating that under fixed prompts and inference settings, the evaluation model behaved as a stable measurement instrument at the transcript level.^15^ In the human validation corpus, human raters reinforced the direction of the primary contrast (high CTF > low CTF) for MITI and key relational measures, but agreement between LLM-based ratings and human scores varied across outcomes. These patterns align with broader concerns that LLM-based evaluators must be validated for each instrument and use case and require ongoing calibration, uncertainty quantification, and transparency regarding its failure modes.^17,19^

Taken together, our findings support LLM-based psychotherapy simulation as a potential versatile, scalable, reproducible and ethically tractable experimental framework for psychotherapy process research. Analogous to *in silico* approaches in other biomedical domains,^4,5^ such a platform can support controlled manipulations of therapist behaviours and patient characteristics that are difficult to randomise in vivo; enable complex multifactorial designs with balanced sample distributions even with rare or notoriously underrepresented participant features; generate large, balanced corpora for method development (e.g., automated fidelity or integrity assessment); and help generate, prune, and prioritise mechanistic hypotheses. In this sense, *in silico* psychotherapy may be most usefully conceptualised as a preclinical research stage: a testbed for probing process theories and measurement strategies and for designing and optimising innovative intervention strategies before advancing to resource-intensive studies with human participants.^17,19,20^

At the same time, the present results also outline clear boundary conditions. The credibility gap relative to real transcripts and the outcome-dependent validity of LLM-based evaluations underline that this platform should not be treated as a substitute for clinical evidence derived from patient outcomes.

### Strengths, limitations and generalisability

Our study has multiple strengths. First, it employs an innovative simulation-based experimental psychotherapy research framework leveraging locally deployed performant open-weight LLMs under reproducible conditions. Second, it performs a controlled manipulation of common therapeutic factors, which are of high theoretical and clinical interest but are challenging to study in conventional experimental research settings. Third, it implements multiple manipulation checks, benchmarking against human therapy sessions, and human evaluation subsample validations to facilitate appropriate interpretation of the results.

Despite these strengths, the study has several limitations that delineate the next steps. First and foremost, while LLMs can simulate remarkably human-like therapeutic dialogues, they remain imperfect models of human psychotherapy participants, exhibiting structural and behavioural deviations that limit the real-world applicability of the findings. In this proof-of-concept implementation, the LLM-based models relied primarily on the opaque statistical associations stored in the model weights. More complex and psychologically more explicit model architectures may contribute to mitigating the substantial empirical credibility and text structure differences between simulated and real sessions.^37^ Improving turn-taking realism, controlling verbosity, and modelling conversational repair (e.g., hesitations, interruptions, partial answers) may be particularly critical for applications where human-likeness is a primary requirement. Second, both the simulating and evaluating LLMs may carry theoretically or clinically relevant biases. These may systematically distort simulated behaviour or its evaluation, undermining the generalisability and clinical interpretability of the findings, particularly in populations underrepresented in the training corpora^20,21^. Notably, shared bias between the simulating and evaluating LLMs may also introduce evaluation “blind spots”, posing a structural limitation on the validity of LLM-based ratings. The observed outcome-dependent agreement between LLM and human ratings may reflect not only model bias but also small size or insufficient MI-specific training of the human raters. Nevertheless, bias-auditing, human benchmarking, outcome-specific calibration, and reporting of uncertainty remain essential when interpreting simulation-based results.^17,19^ Third, this proof-of-concept study relies on a single therapeutic approach (MI), a limited variety of patient demographics and clinical presentations, a single base model for all sessions and simulated participants, and a fixed prompting strategy. The applicability of the *in silico* framework to other patient populations, therapeutic approaches, foundational models, and prompting or decoding configurations remains unknown, and warrants explicit testing, ideally using shared benchmarks and ablation studies. Fourth, in this proof-of-concept study, the simulations were limited to a single written format session, omitting the influence of non-verbal behaviour, prosody, or session-to-session dynamics that are central to psychotherapy. Outcome metrics were also restricted to observation-based evaluations. Critically, “patient changes” could not be measured through self-reported metrics analogous to testing real individuals and should be interpreted as process proximal indicators reflecting stance and commitment language, not as clinical effects. Extending simulations to longitudinal and multimodal formats and developing more adequate evaluation strategies is an important but technically challenging direction. And fifth, unlike the simulations, the LLM evaluations relied on a proprietary model, potentially challenging future reproducibility.

### Future directions

Future *in silico* psychotherapy research should therefore expand the diversity of patient characteristics, clinical presentations, therapeutic approaches, therapist styles, languages, and care settings; extend simulations to longitudinal and, where feasible, multimodal formats; systematically map which manipulations produce robust changes across therapist integrity, dyadic process, linguistic features, and evaluator agreement; and more precisely delineate boundary conditions, including the circumstances under which simulations fail, become implausible, or lose measurement validity. Further, a systematic exploration of more sophisticated simulation frameworks integrating explicit representations of internal cognitive, emotional or motivational states, paralleling deliberative agent architectures and belief–desire–intention (BDI) type models^38,39^, may enhance simulation credibility, experimental control over latent psychological variables, and overall ecological validity of the effects observed in simulations. Across these next steps, transparency, ethics, bias auditing, adversarial testing, as well as bench-marking against human judgement and real world materials should be integrated as constitutive elements of study design.^6,17,19–21,40,41^

In the context of LLM-based clinical and training tools, LLM-based simulations have been proposed for model training, fidelity assessment and safety testing purposes.^13^ The present study contributes to ongoing discussions about the role of generative AI in healthcare by suggesting that such systems may also be valuable as instruments for theory-guided methodological research.

## Conclusion

In this preregistered proof-of-concept, fully automated LLM-based motivational interviewing simulations were feasible at scale, showed robust experimental fidelity, and recovered theoretically concordant effects of a therapist-side experimental manipulation of common therapeutic factors. Importantly, effects extended beyond targeted relational process measures such as empathy and alliance to MI-specific process metrics and selected proxy indicators of simulated patient improvement. These findings support *in silico* psychotherapy as a promising, scalable and cost-effective preclinical platform for controlled and reproducible psychotherapy research and method development. However, remaining challenges related to simulation credibility and LLM evaluation alignment underscore that such simulations should complement rather than substitute studies with human participants. With continued validation, human benchmarking, transparency, and systematic bias-aware evaluation, *in silico* psychotherapy research may become a valuable component of the methodological pipeline for psychotherapy science, enabling more rapid, iterative, and theory-driven investigations.^4–7,17,19–21,25,27–29,41^

## Code availability statement

The code and prompts used to generate and evaluate simulations and perform analyses will be made available upon reasonable request to the corresponding author.. Some components rely on third-party LLM APIs and may require appropriate access credentials.

## Author contributions

GM conceived and supervised the study project. GM, HAB and KH developed the methodology. HAB, KH and SL implemented the LLM-based simulations. KH implemented the LLM-based evaluations and the assessment of rule-based outcomes and analysed and interpreted simulation evaluation data. EB and LS prepared and conducted the human evaluations. SB analysed and interpreted human evaluation data. GM and KH wrote the original draft. ZE, MT and IJT contributed to conceptualization and methodological guidance. ZE, RS, MT and IJT contributed to the critical revision of the manuscript. All authors read and approved the final manuscript.

## Funding, Competing Interests and Declarations

This work was funded in part by the Rhineland-Palatinate Research Initiative 2024–2028 (No. FI/P 2026-1b). The funder had no role in study design; in the collection, analysis or interpretation of data; in the writing of the report; or in the decision to submit for publication. The grants listed under competing interests are general author disclosures and were not specific to this study. GM & RS received funding in the context of a Horizon Europe project from the Swiss State Secretariat for Education, Research and Innovation (SERI) under contract number 22.00094. Further, GM & RS received funding from Wings Health Inc. in the context of a proof-of-concept study. GM received funding from the Swiss Heart Foundation under project no. FF21101, from the Research Foundation of the International Psychoanalytic University (IPU) Berlin under projects no. 5087 and 5217, from the Swiss National Science Foundation (SNSF) under project no. 100014_135328, from the German Federal Ministry of Education and Research under budget item 68606, and from the Hasler Foundation under project no. 23004. GM is co-founder and holds stock in Therayou AG, which is active in the field of digital and blended mental healthcare. GM receives royalties from publishing companies as author, including a book published by Springer, and an honorarium from Lundbeck for speaking at a symposium. Furthermore, GM is compensated for providing psychotherapy to patients, acting as a supervisor, serving as a self-experience facilitator (‘Selbsterfahrungsleiter’), and for postgraduate training of psychotherapists and supervisors. RS received a speaker honorarium from Novartis. RS is chairman of the Basel Institute for Psychosomatic Medicine (BIPM) and founder and managing director of the Psychosomatic and Psychosocial Services GmbH, that develops and implements psychosomatic and psychosocial training and continuing education programs. He is a member of the board of trustees of the Foundation Psychosomatic and Social Medicine (Ascona Foundation). KH co-founded the inactive startup RevealApp dedicated to NLP-based content search in unstructured text data. The authors declare no other potential conflicts of interests. The research activities were fully independent, and there were no intellectual or financial proprietary claims.

We used AI-based tools, including ChatGPT and OpenEvidence to support manuscript preparation. Further, we used publicly available search technologies, which we recognize likely utilise AI capabilities. We confirm that the contributions of AI were strictly in an assistive capacity. AI was not involved in conceptual tasks. Human oversight was continuously employed to ensure the accuracy of content and address any ethical concerns.

## Data Availability

All data produced in the present study are available upon reasonable request to the authors.

No new data were collected from human patients. Derived outcome data underlying the figures and tables (transcript-level ratings and text-derived metrics for simulated sessions) are available upon reasonable request to the corresponding author. The real-world MI transcripts used as comparators were obtained from publicly accessible third-party sources (see also Supplementary Methods 1.10).

