## Supplement for "Simulating Psychotherapeutic Sessions with Generative Artificial Intelligence: A Proof-of-Concept Study of *In Silico* Psychotherapy Research"


Katalin Horváth[a,b,c], Shiv Longani[a], Shir Brill[b,c], Harry A. Ballantyne[b,c,d], Rainer Schaefert[c], Marion Tegethoff[e], Liam Schirmer[a], Emma Börner[a], Ingo J. Timm[f,g], Zohar Elyoseph[g,h], Gunther Meinlschmidt[a,b,c*]

a. Clinical Psychology and Psychotherapy – Methods and Approaches, Department of Psychology, Trier University, Trier, Germany.
b. Department of Digital and Blended Psychosomatics and Psychotherapy, Psychosomatic Medicine, University Hospital and University of Basel, Basel, Switzerland.
c. Department of Psychosomatic Medicine, University Hospital and University of Basel, Basel, Switzerland.
d. Department of Ophthalmology, University Hospital Zurich and University of Zurich, Zurich, Switzerland
e. Department of Health Psychology, Institute of Psychology, RWTH Aachen, Aachen, Germany.
f. Business Informatics I, Department IV, Trier University, Trier, Germany.
g. German Research Center for Artificial Intelligence (DFKI). Research Department Cognitive Social Simulation, Trier, Germany.
h. Faculty of Education, School of Therapy Counselling and Human Development, University of Haifa, Haifa, Israel.

*** Corresponding Author:**

Prof. Dr. Gunther Meinlschmidt, Trier University, Universitaetsring 15, 54296 Trier, Germany; University Hospital Basel, Hebelstr. 2, 4031 Basel, Switzerland; Phone: +49 651 201 1999; +41 61 328 46 33, E-Mail:

**Supplementary Materials SM1**

Technical implementation, inference parameters, and quality-control rules for *in silico* session generation.

—

This supplement describes prompt modularisation and execution logic in detail and summarises all decoding and run-time parameters required to reproduce the simulations. Full prompt templates are available from the authors on reasonable request.

#### *SM1.1 Computing environment and model deployment*

All simulations were batch-executed on the SciCORE+ secure high-performance computing (HPC) infrastructure at the University of Basel on the 25th and 26th of April 2025. Simulations were executed using linked NVIDIA A100 GPUs (40 GB each) as independent, non-interactive batch jobs using standard HPC scheduling. Dialogues were generated using a locally deployed open-weight conversational large language model (Llama-3.3-70B-Instruct), ensuring that no data were transmitted to external services during inference. The model was run with 8-bit quantisation to reduce memory requirements and to enable efficient inference on the available hardware.

Model inference was performed using llama.cpp, accessed via the Python interface llama-cpp-python. Locally run python scripts orchestrated message passing between LLM-based participants, quality control, logging, and protocol assembly. All generated transcripts, associated run metadata, and intermediate model outputs were automatically retained as part of the run logs. Analyses reported here were based exclusively on the final generated transcripts.

#### *SM1.2 Simulation orchestration and LLM-based architecture*

We generated each simulated MI session by orchestrating multiple role-specific LLM-based entities, all instantiated from the same base model and differentiated solely by role prompts and role-specific constraints:

1. LLM-based therapist (MI delivery): Generated therapist utterances consistent with MI and the assigned therapist persona. The experimental manipulation (high vs low common therapeutic factors, CTF) was implemented exclusively through an additive prompt module for this entity.
2. LLM-based patient (vignette-constrained patient): Generated patient utterances consistent with the assigned patient vignette and patient persona.
3. LLM-based interjections (therapist backchannels): At randomly selected points in the session, short therapist backchannels/acknowledgements were generated, intended to mimic active listening and improve conversational naturalness.
4. LLM-based monitoring (closure decision): After each exchange, this entity evaluated whether the dialogue had reached a plausible closure and should end, enabling adaptive termination rather than enforcing a fixed number of turns.

A single session was produced by iterative turn-taking between LLM-based patients and therapists, with optional insertion of interjections and repeated monitoring for termination.

#### *SM1.3 Prompt modularisation, role separation and role personas*

We constructed prompts as modular templates, combined at runtime to create an "effective" system context for each LLM call. Across all LLM-based entities, prompts enforced a transcript-style output (single-speaker utterances in plain spoken language) and prohibited meta-commentary, stage directions, and list formatting.

For the LLM-based therapist, the effective prompt comprised:

- A base therapist role module (identity constraints; "speak only as therapist"; transcript style constraints).
- A MI scaffold module (MI process framing and behavioural constraints consistent with MI practice).
- A therapist persona module (persona-specific characteristics held constant across conditions).
- A CTF condition module (high vs low CTF), added identically across all sessions in a given condition.
- An alternative version with a backchanneling module, ensuring that the therapist occasionally signals active listening. For technical reasons, this alternative version also triggered a corresponding truncation and re-generation of the original patient utterance during which the backchanneling occurred.

For the LLM-based patient, the effective prompt comprised:

- A base patient role module (identity constraints; "speak only as patient"; transcript style constraints).
- A patient vignette/persona module (case-specific contextual information held constant across conditions).

For the LLM-based interjections, prompts constrained outputs to brief backchannels/acknowledgements without narrative elements. For the LLM-based monitoring, prompts constrained outputs to a structured decision about whether the session should terminate, based on whether a plausible MI session end point had been reached.

Prompt templates used for session generation are available on request from the corresponding author.

*Patient personas*

Eight standardised vignettes for the patient personas were constructed prior to simulation, each representing an adult individual presenting with a behavioural or mental health-related concern amenable to MI (e.g., ambivalence regarding change). Vignettes were derived from publicly available MI transcripts and were written to include a coherent presenting problem, relevant contextual details, and plausible expressions of ambivalence. Vignette texts were used exclusively as prompts to initialise the LLM-based patient and were held constant across conditions. We provide a summary of the patient vignettes in Supplementary Table ST1.

*Therapist personas*

Eight therapist personas were developed to introduce controlled heterogeneity in non-manipulated therapist characteristics (e.g., communication style descriptors or further therapeutic expertise besides MI), while maintaining MI as the implemented therapeutic approach across all personas. Personas were orthogonal to the experimental manipulation: the CTF manipulation was implemented by condition-specific instruction modifications added to the LLM-based therapist prompts, whereas all other persona and role instructions were identical across conditions. Potential persona-dependent baseline CTF characteristics were accounted for through the fully crossed design and through random intercepts in the mixed effects model analyses. Full therapist persona characteristics are summarised in Supplementary Table ST2.

#### *SM1.4 Inference settings*

All decoding and inference parameters were prespecified and held constant within each LLM-based role across all sessions, to ensure that between-condition differences were attributable to the CTF prompt manipulation rather than to sampling hyperparameters. We used role-specific inference settings to balance coherence and variability across therapist and patient text and to constrain the interjection and monitoring roles.

The decoding and inference parameters used for each LLM-based simulation component (fixed across sessions within LLM-based component) were as follows:

- Backend/model:
    - Model: Llama-3.3-70B-Instruct
    - Quantisation: 8-bit (Q8_0)
    - Context window (n_ctx): 40000
- Sampling/decoding:
    - Temperature: 0.95 for all LLM queries
    - Top-p: 1.0 (meaning disabled)
    - Top-k: 0 (meaning disabled)
    - Minimum probability: 0.02
    - Typical-p / TFS / Mirostat: not used
    - Repetition penalty and/or frequency/presence penalties: 1 (meaning no penalty)
- Output control:
    - Maximum tokens per turn (max_tokens): 1000 for all LLM queries
    - Stop sequences: none specified
- Determinism/randomness:

  - RNG seed handling: set per call for Therapist, Patient and Interjection and per run for Monitoring, using the Python library "secrets".
- Performance-related:
  - GPU offload / tensor-parallel settings: all 80 layers
  - Batch size / threads (if set explicitly): 4

#### *SM1.5 Dialogue loop and termination criteria*

*Turn-taking and context handling*

The transcript was generated as an ordered sequence of alternating therapist and patient utterances. At each step, the LLM was provided with the session context required to maintain coherence (i.e., the dialogue history accumulated so far) together with the role-specific system context.

*Termination via LLM-based monitoring*

After each exchange, the LLM-based monitoring evaluated whether the session had reached a plausible end point (e.g., a closing summary and next-step planning consistent with an MI session). The simulation terminated when closure criteria were met.

*Termination via technical time restrictions*

Computational resources were assigned to simulation jobs for the requested pre-defined amount of time (1.5 hours) through the HPC scheduler. If the LLM-based monitoring had not triggered termination before the resource reservation expired, the simulation process eventually got interrupted without saving a final transcript, and the session was recorded as a technical failure in feasibility summaries. Each simulation job was attempted exactly once.

#### *SM1.6 Rule-based runtime validity checks and regeneration logic*

To reduce common artefacts in LLM-generated transcripts, we applied a rule-based validity filter to each candidate utterance *before* appending it to the transcript and forwarding it to the counterpart LLM-based entity. When an utterance failed the validity filter, it was rejected and a replacement was generated using the same LLM role and the same conversation context.

We triggered regeneration when outputs contained prespecified character patterns potentially indicating any of the following violations:

- Explicit list formatting, including bullet points or numbered lists (multiple newlines starting with numbers or bullet point characters).
- Narrative descriptions of actions or non-verbal behaviour (e.g., stage directions), rather than spoken dialogue (separating characters for the descriptions such as "(" or "*").
- Repetition of earlier content suggesting loss of progression (an output matching the previous utterance of the participant).

There was no pre-specified boundary to the number of regeneration attempts beyond the simulation-level technical time limitation enforced by the HPC job scheduler.

#### *SM1.7 Randomisation, batching, and run bookkeeping*

*Simulation design instantiation*

We implemented the fully crossed design (8 patient vignettes × 8 therapist personas × 2 CTF conditions × 4 replicates per combination) using the combination of the four indices to consistently and unambiguously identify simulation job, output and log/error files, generated session transcript files, and data records. The simulation script was identical across all runs and received the experimental condition, therapist persona, patient persona, and repetition index as command-line arguments.

*Run-order randomisation and batching*

Simulations were submitted to the HPC job scheduler in an explicitly randomized order to minimise serial artefacts during the batch run (e.g., due to resource heterogeneity); the randomized sequence was generated at runtime prior to submission and was logged for record-keeping.

*Logging and metadata*

For each attempted run, we retained:

- Run identifiers (patient ID, therapist ID, condition, replicate).
- The standard output file of the simulation script containing job identifiers, random seeds, all LLM interactions with prompts and outputs, and the results of cleaning and quality control steps including invalid-output rejections and regeneration attempts.
- The error file of the simulation script logging diagnostic information, especially from third party software.
- Job metadata from the HPC cluster.
- The final cleaned transcript as a text file if the simulation was successful. For each session, three aligned protocol representations were stored:
    - 1. Full dialogue protocol: interleaved therapist and patient utterances in chronological order (used for MI fidelity and process ratings, turn-taking, and credibility judgements).
    - 2. Therapist-only protocol: concatenated therapist utterances (used for therapist-specific linguistic metrics).
    - 3. Patient-only protocol: concatenated patient utterances (used for patient-specific linguistic metrics and proxy patient improvement metrics).

Failed runs did not produce simulation transcript files and were identified post hoc via missing output files for expected identifier combinations.

The standard output and error logs as well as the job metadata were retained for diagnostic and audit purposes but were not parsed or used in any downstream analyses. All further analyses were conducted exclusively on the final simulation transcript files produced by successfully completed runs.

#### *SM1.8 Alternative statistical models and sensitivity analyses*

To evaluate the effect of the CTF condition, we tested alternative mixed effects models with random effect structures allowing for additional random slopes for therapist persona groups. These yielded very similar main effects and confidence intervals for the CTF condition across all inspected outcome variables, while producing more singular fits and convergence failures for random effects.

For all mixed-effects models, we checked residual diagnostics. Residual values diverged substantially from normality assumptions; hence, we investigated alternative models such as beta regression or robust regression models. These models also resulted in similar main effect patterns across all variables. Further, we compared results to conventional two-sample comparisons (Welch t-tests and Mann–Whitney U tests) as descriptive checks and found patterns consistent with the reported mixed effects models results.

We assessed the potential effect of differential attrition through post-simulation exclusion criteria by re-running all mixed-effects models including all successful simulations. These models produced very similar estimates and CIs to the original analyses.

We also investigated whether CTF effects remain supported after controlling for output length characteristics (mean patient and therapist utterance lengths and total utterance count), since these were affected by the CTF manipulation and could have produced evaluation artefacts. Mixed effects model CTF effects for all MITI global scores, WAI scores and TES scores persisted with slightly decreased estimates and broadened confidence intervals. Simulated observer-inferred proxy patient changes increased, supporting CTF-dependent differences in both importance and confidence ruler improvement across simulations with similar utterance lengths and count. Therapist credibility differences disappeared, while simulated patients in the high CTF condition were rated as less credible when length characteristics were similar. This analysis indicates that most CTF effects in the main analysis remained robust to adjustment for the measured CTF-dependent conversation structure differences, with the exception of therapist credibility. Interestingly, the patient-side estimate changes are consistent with a suppression pattern, suggesting that length-related characteristics may have partly counteracted rather than produced the observed CTF-dependent differences, potentially through their association with perceived patient credibility or attitudes toward the discussed behavioural change.

Further, we estimated multiplicative effects for all quantitative text metrics using log-scaled versions of the mixed effects models, relying on the same fixed and random effects structure:

$$log(Outcome) \sim CTF + (1 \mid patient\ vignette) + (1 \mid therapist\ persona).$$

Since length effects followed very similar patterns across measurement units (sentences, words, syllables and characters), we only included mean word counts per utterance, total word counts, and total utterance counts in the main manuscript.

Specific details and model parameter estimations for these models are available from the authors on reasonable request.

#### *SM1.9 Analysis software*

We used tidyverse for data management; lme4, broom.mixed and lmerTest for mixed-effects models; effectsize, rstatix and boot for effect sizes and non-parametric tests; performance for diagnostics; robustlmm and glmmTMB for sensitivity models; and psych, irr, car, krippendorff, and BlandAltmanLeh for reliability and agreement analyses.

#### *SM1.10 LLM-based evaluations*

MITI 4.2 global ratings, patient change rulers, credibility ratings, TES, WAI-S-O and severe simulation errors were evaluated using GPT-5 via scripted API calls after completion of all simulations. For each outcome, the evaluation prompt included (i) a concise description of the instrument, (ii) scoring anchors and response format requirements, and (iii) the transcript to be rated. For the importance and confidence scores, the transcript contained the patient utterances only. All other metrics were scored based on the full transcript version. The evaluation model was not provided with the simulation condition or any metadata indicating whether a transcript was simulated or real. Rating tasks were conducted independently; the model was not informed about other outcomes or prior ratings for the same transcript.

To improve robustness, each transcript was rated five times per outcome using identical instructions and fixed inference settings; scores were aggregated at the transcript level (mean across repeats). Automated extraction of numeric scores from the model's output was implemented with a predefined parsing routine built into the LLM evaluation pipeline and included a rule-based validity control step allowing for up to five re-generations of the entire evaluation round in case of technical errors or format deviations. To further mitigate extraction errors, outputs were audited for adherence to the expected format; when deviations occurred, values were manually recovered when unambiguous or coded as missing when not recoverable.

LLM-based evaluations were executed via the OpenAI batch API in six batches, each containing all queries belonging to the same evaluation criteria (Simulation errors, MITI, WAI, TES, Credibility and Rulers, respectively.). The five measurement repetitions per evaluated session were included in the same batch. The processing pipeline included a rule-based formal validity assessment of the extracted scores. Queries that failed on the server or failed to produce a formally valid score for any other reason were repeated in up to five subsequent batch queries as planned. Due to repetitions and asynchronous batch processing, requests were executed between 02.09.2025 and 04.09.2025. The resolved model version reported in the outputs was gpt-5-2025-08-07. No missing or invalid scores were identified after the process was finished.

The custom credibility scale was assessed with the following wording:

"Evaluate whether the "Therapist" and the "Patient" participants in the provided protocol are more likely humans or more likely simulations, as a score between 0 (certainly a human) and 10 (certainly a simulation)." (Scores were inverted through the analysis, corresponding to the identical but inverted human ruler evaluation scales).

The custom importance and confidence ruler scales were assessed with the following wording, in four separate items (importance/confidence x before/after):

"How *IMPORTANT/CONFIDENDT* do you think this change was for "Patient" just *before/after* the session, on a scale of 1 (not important at all) to 10 (very important)?"

#### ***SM1.11 Original Motivation Interviewing Transcripts***

The real-world MI transcripts used as comparators were obtained from the following publicly accessible third-party sources:

https://casaa.unm.edu/tools/miti.html

https://attcnetwork.org/products_and_resources/motivational-interviewing-assessment-supervisory-tools-for-enhancing-proficiency-miastep/

Martino, S., Ball, S.A., Gallon, S.L., Hall, D., Garcia, M., Ceperich, S., Farentinos, C., Hamilton, J., and Hausotter, W. (2006) Motivational Interviewing Assessment: Supervisory Tools for Enhancing Proficiency. Salem, OR: Northwest Frontier Addiction Technology Transfer Center, Oregon Health and Science University.

Transcripts no. 7 and no. 8 were published in Spanish and were translated using GPT 4 via the OpenAI ChatGPT browser interface in August 2024.

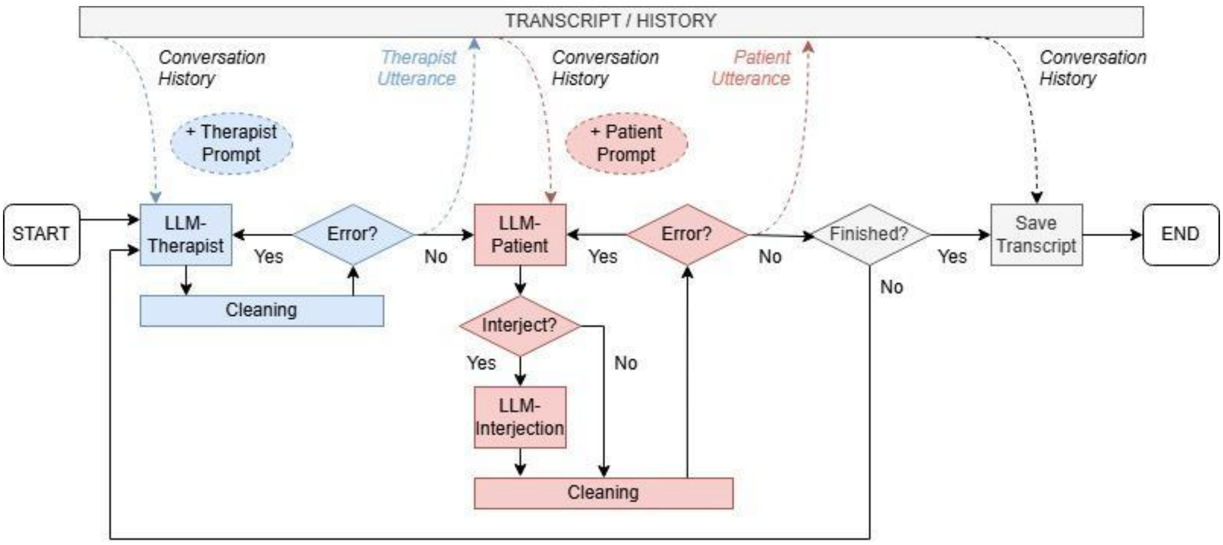


### Supplementary Figure SF1

Simulation process flowchart.

*Notes: Flowchart of the simulation pipeline, including message passing between LLM-based therapists and patients, insertion of brief interjections (backchanneling), monitoring for plausible session closure, and rule-based output validity checks (with regeneration when criteria were violated). Post hoc screening for severe simulation errors and the resulting analysis sets are described separately in Figure 2. Abbreviations: LLM, large language model.*

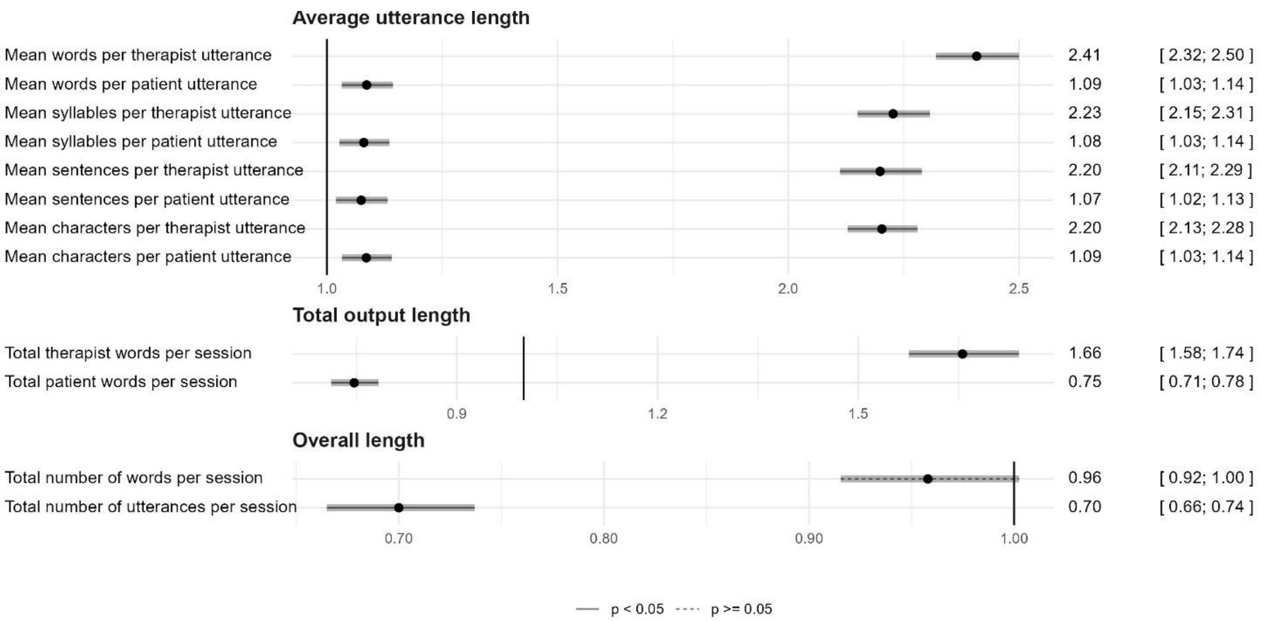


**Supplementary Figure SF2**

Sensitivity analysis: multiplicative effects for transcript-level text metrics.

*Notes: Sensitivity analysis for transcript-derived text metrics using log-linear mixed-effects models. Points show multiplicative High/Low CTF ratios (with 95% CIs). Ratios >1 indicate larger values in the high-CTF condition. Abbreviations: CTF, common therapeutic factors; low CTF: low levels of common therapeutic factors (CTF); high CTF: high levels of CTF*

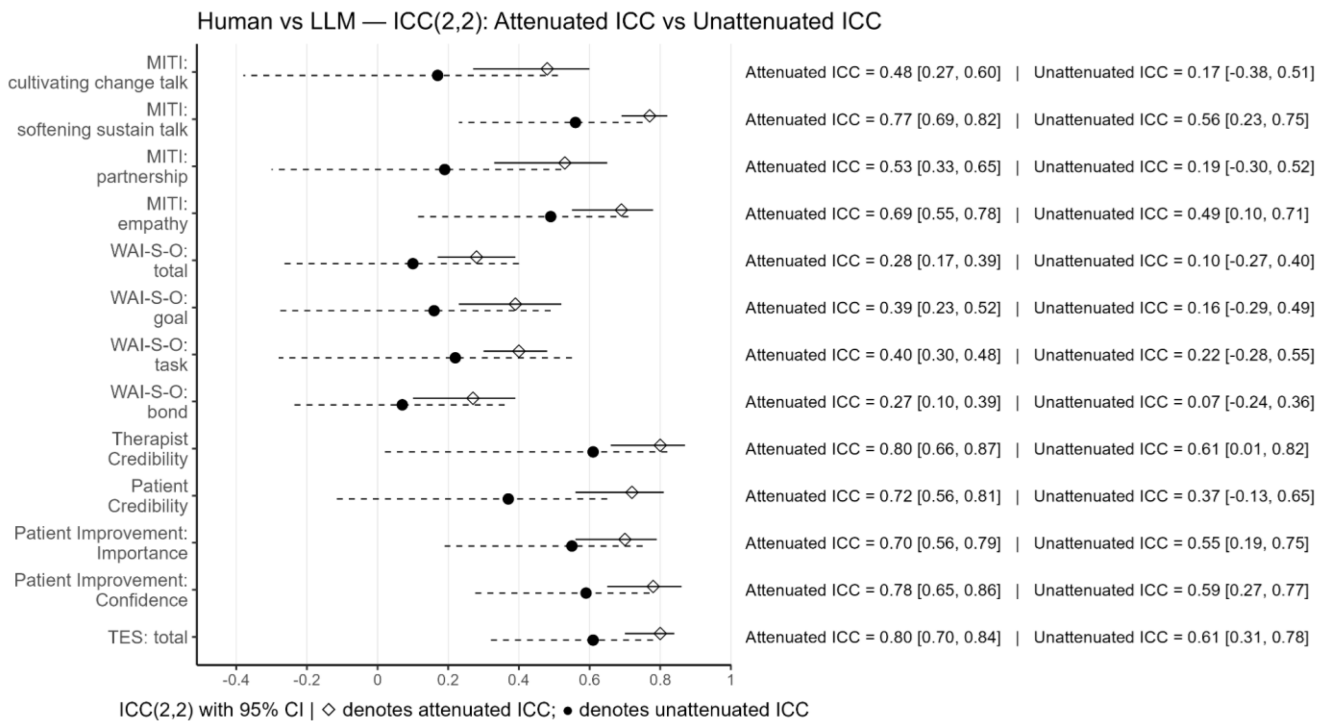


### Supplementary Figure SF3

Human–LLM agreement for outcome ratings.

*Notes: Agreement between GPT-5-based ratings and blinded human ratings for outcomes in the validation subset. Points show ICC(2,2) estimates with 95% CIs. “Unattenuated” (uncorrected) ICCs reflect observed agreement; attenuation-corrected (“attenuated”) ICCs adjust for within-rater internal consistency. Interpretation: poor < 0.40, fair 0.40-0.59, good 0.60-0.75, excellent > 0.75. Abbreviations: LLM, large language model; ICC, intraclass correlation coefficient; MITI, Motivational Interviewing Treatment Integrity; WAI-S-O, Working Alliance Inventory-Short Observer form; ES, Therapist Empathy Scale*

**Supplementary Table ST1**

Patient vignettes used to initialize LLM-based patients.

| | Patient Sociodemographics | | Clinical Information | Patient's Psychosocial Context | | | Vignette Length Metrics | | |
|---|---|---|---|---|---|---|---|---|---|
| **#** | **Age (years)** | **Gender** | **Presenting Problem(s)** | **Stressors and Life Events** | **Coping Mechanisms and Resilience** | **Socio-economic Background** | **No. of Sentences** | **No. of Words** | **No. of Characters** |
| 1 | 53 | Male | Smoking addiction | Social stigma, financial cost, identity conflict | Autonomy, self-expression | NA | 6 | 185 | 1,294 |
| 2 | 40 | Male | Smoking addiction | Health/performance impact, financial cost | Athletic identity, autonomy | NA | 7 | 150 | 1,064 |
| 3 | 48 | Female | Low motivation for exercise, wants to improve health, diabetes management | Health concerns, stress about exercising | Disciplined dietary control, high conscientiousness, values health | Small-town context | 8 | 176 | 1,207 |
| 4 | 38 | Male | Parenting difficulties | Incident with social worker, conflicts about respect and authority | Commitment to be a better father, considering new parenting methods, seeking psychotherapy | Blue-collar identity, suburban context | 10 | 220 | 1,511 |
| 5 | 56 | Male | Stress and drinking issues | Legal issues, family strains, employment risk, multiple life events | Independence, social network | Rural town, drinking culture | 5 | 149 | 1,049 |

| | | | | | | | | | |
|---|---|---|---|---|---|---|---|---|---|
| 6 | 34 | Male | Stress and drinking issues | Legal issues, job risk, performance pressures, social pressure | Commitment to career | NA | 10 | 221 | 1,509 |
| 7 | 28 | Female | Stress | Separation from a partner, custody concerns, societal judgment, pressure from authorities | Active social life, dedication to job and children | Suburban context, stigma | 9 | 190 | 1,321 |
| *8* | 28 | Female | Stress, occasional cocaine use | Single motherhood, societal judgment, sadness and loneliness, custody concerns, substance use | Prioritizing daughters' well-being | Mid-sized town context, stigma around substance use | 7 | 163 | 1,140 |

*Notes: Summary characteristics of the eight standardized fictional patient vignettes used to initialize LLM-based patients (demographics, presenting problem, ambivalence/change talk, and key contextual stressors). Vignettes were constructed to represent MI-amenable presentations. Full vignette texts were held constant across conditions and are available on request. Abbreviations: MI, motivational interviewing; NA, not assessed.*

### Supplementary Table ST2

Therapist personas used to initialize LLM-based therapists.

| # | Name | Gender | Ethnicity | Clinical Experience | Self-Awareness | Cultural Sensitivity | Inclusivity | Personal Practices | Professional Engagement | Interpersonal Skills | Therapeutic Background |
|---|---|---|---|---|---|---|---|---|---|---|---|
| 1 | Emily | Female | Asian-American | Over 10 years | High, maintains healthy boundaries and awareness of own influence | Sensitive to cultural, ethnic, and gender influences on mental health | Committed to creating an inclusive, affirming space for all clients | Mindfulness meditation for emotional balance | Connects with colleagues at conferences, avid reader | Exceptional, prioritizes building strong therapeutic alliances | Evidence-based, tailored to individual client needs |
| 2 | Michael | Male | Caucasian | 5 years | Moderate, engages in regular self-reflection | Aware of cultural differences, seeks to learn more | Open to working with diverse populations | Regular exercise and time in nature | Active in professional organizations | Good, focuses on active listening and empathy | Cognitive-Behavioral Therapy (CBT) |
| 3 | Sarah | Female | African-American | 15 years | Very high, prioritizes self-care and personal growth | Highly attuned to racial and cultural trauma | Passionate advocate for social justice and equity | Journaling and creative self-expression | Volunteers at community mental health clinic | Excellent, uses humor and warmth to connect with clients | Eclectic, drawing from various modalities |
| 4 | Liam | Male | Irish-American | 20 years | High, seeks consultation and supervision when needed | Respectful of cultural beliefs about mental health and treatment | Strives to create a welcoming environment for all | Maintains work-life balance and hobbies | Collaborates with other healthcare providers | Very good, maintains professional boundaries while being approachable | Psycho-pharmacology and supportive therapy |
| 5 | Priya | Female | Indian-American | 8 years | High, actively explores own biases and cultural identity | Integrates cultural values and practices into therapy | Affirming of diverse sexual orientations and gender identities | Yoga and spirituality | Conducts research on multicultural therapy | Strong, adapts communication style to client needs | Multicultural and feminist therapy |
| 6 | David | Male | Chinese-American | 12 years | Moderate, balances personal and professional life | Sensitive to family dynamics and cultural norms | Inclusive of non-traditional family structures | Family time and community involvement | Presents at workshops on couples therapy | Exceptional, creates a safe and supportive environment | Systemic and solution-focused therapy |

| | | | | | | | | | | | |
|---|---|---|---|---|---|---|---|---|---|---|---|
| 7 | Kim | Non-binary | Asian-American | 25 years | Exceptionally high, undergoes personal analysis | Understands the impact of acculturation and immigration | Welcomes clients from all socioeconomic backgrounds | Engages in personal therapy | Supervises and mentors junior colleagues | Excellent, uses insight and interpretation to deepen understanding | Psychodynamic therapy |
| 8 | Alex | Non-binary | Mixed-race | 7 years | High, uses art-making for self-reflection | Affirming of diverse identities and experiences | Creates a safe space for LGBTQ+ clients | Art-making and self-care | Facilitates art therapy groups in the community | Warm, uses creativity to facilitate self-expression | Expressive arts therapy |

*Notes: Summary characteristics of the eight therapist personas used to initialize LLM-based therapists (Gender, race/ethnicity, clinical experience, self-awareness, cultural sensitivity and inclusivity, personal practices, professional engagement, interpersonal skills, and therapeutic approach). Abbreviations: LLM, large language model; LGBTQ+, lesbian, gay, bisexual, transgender, queer, and related identities; CBT, cognitive behavioral therapy. Full persona narratives are available on request*

**Supplementary table ST3**

Distribution of all manipulated predictor variables across attempted, successful and included simulations.

| Variable levels | Simulation runs | | Succeeded | | Included | |
|---|---|---|---|---|---|---|
| | ***n*** | **%** | ***n*** | **%** | ***n*** | **%** |
| *CTF condition* | *512* | | *506* | | *480* | |
| low CTF | 256 | 50.00 | 250 | 49.41 | 237 | 49.38 |
| high CTF | 256 | 50.00 | 256 | 50.59 | 243 | 50.62 |
| *Patient ID* | *512* | | *506* | | *480* | |
| 1 | 64 | 12.50 | 63 | 12.45 | 57 | 11.88 |
| 2 | 64 | 12.50 | 64 | 12.65 | 63 | 13.12 |
| 3 | 64 | 12.50 | 64 | 12.65 | 60 | 12.50 |
| 4 | 64 | 12.50 | 64 | 12.65 | 59 | 12.29 |
| 5 | 64 | 12.50 | 63 | 12.45 | 61 | 12.71 |
| 6 | 64 | 12.50 | 62 | 12.25 | 57 | 11.88 |
| 7 | 64 | 12.50 | 62 | 12.25 | 60 | 12.50 |
| 8 | 64 | 12.50 | 64 | 12.65 | 63 | 13.12 |
| *Therapist ID* | *512* | | *506* | | *480* | |
| 1 | 64 | 12.50 | 64 | 12.65 | 60 | 12.50 |
| 2 | 64 | 12.50 | 63 | 12.45 | 58 | 12.08 |
| 3 | 64 | 12.50 | 64 | 12.65 | 64 | 13.33 |
| 4 | 64 | 12.50 | 64 | 12.65 | 62 | 12.92 |
| 5 | 64 | 12.50 | 64 | 12.65 | 58 | 12.08 |
| 6 | 64 | 12.50 | 62 | 12.25 | 60 | 12.50 |
| 7 | 64 | 12.50 | 62 | 12.25 | 56 | 11.67 |
| 8 | 64 | 12.50 | 63 | 12.45 | 62 | 12.92 |

*Notes: Frequencies of CTF conditions, patient vignette IDs, and therapist persona IDs across initiated runs (N=512), technically successful session generations (N=506), and included sessions after post hoc QC exclusions (N=497). Abbreviations: CTF, common therapeutic factors; low CTF, low levels of common therapeutic factors (CTF); high CTF, high levels of CTF. for a detailed list of therapist and patient profiles see the online supplement.*

**Supplementary Table ST4**

Fitted Mixed Effects Model parameters.

| **Outcomes** | **Effect type** | **Group** | **Term** | **Estimate** | ***SE*** | ***p*** | **95% CI** |
|---|---|---|---|---|---|---|---|
| MITI: cultivating change talk | Fixed | | Intercept | 3.76 | .032* | 0.00 | [3.70, 3.83] |
| MITI: cultivating change talk | Fixed | | CTF condition | 0.54 | .030* | 0.00 | [0.48, 0.60] |
| MITI: cultivating change talk | Random | PatientID | sd Intercept | 0.07 | | | |
| MITI: cultivating change talk | Random | TherapistID | sd Intercept | 0.00 | | | |
| MITI: cultivating change talk | Random | Residual | sd Observation | 0.33 | | | |
| MITI: softening sustain talk | Fixed | | Intercept | 3.16 | .060 | 0.00 | [3.03, 3.29] |
| MITI: softening sustain talk | Fixed | | CTF condition | 0.95 | .046* | 0.00 | [0.86, 1.04] |
| MITI: softening sustain talk | Random | PatientID | sd Intercept | 0.14 | | | |
| MITI: softening sustain talk | Random | TherapistID | sd Intercept | 0.00 | | | |
| MITI: softening sustain talk | Random | Residual | sd Observation | 0.50 | | | |
| MITI: partnership | Fixed | | Intercept | 3.81 | .031* | 0.00 | [3.74, 3.88] |
| MITI: partnership | Fixed | | CTF condition | 0.86 | .030* | 0.00 | [0.80, 0.92] |
| MITI: partnership | Random | PatientID | sd Intercept | 0.06 | | | |
| MITI: partnership | Random | TherapistID | sd Intercept | 0.02 | | | |
| MITI: partnership | Random | Residual | sd Observation | 0.33 | | | |
| MITI: empathy | Fixed | | Intercept | 3.48 | .035* | 0.00 | [3.40, 3.55] |
| MITI: empathy | Fixed | | CTF condition | 0.75 | .032* | 0.00 | [0.69, 0.82] |
| MITI: empathy | Random | PatientID | sd Intercept | 0.06 | | | |
| MITI: empathy | Random | TherapistID | sd Intercept | 0.04 | | | |
| MITI: empathy | Random | Residual | sd Observation | 0.35 | | | |
| WAI-S-O: total | Fixed | | Intercept | 60.17 | .374 | 0.00 | [59.34, 61.00] |

| | | | | | | | |
|---|---|---|---|---|---|---|---|
| WAI-S-O: total | Fixed | | CTF condition | 3.14 | .251 | 0.00 | [2.65, 3.63] |
| WAI-S-O: total | Random | PatientID | sd Intercept | 0.93 | | | |
| WAI-S-O: total | Random | TherapistID | sd Intercept | 0.00 | | | |
| WAI-S-O: total | Random | Residual | sd Observation | 2.74 | | | |
| WAI-S-O: goal | Fixed | | Intercept | 19.76 | .199 | 0.00 | [19.32, 20.21] |
| WAI-S-O: goal | Fixed | | CTF condition | 0.07 | .116 | 0.56 | [-0.16, 0.29] |
| WAI-S-O: goal | Random | PatientID | sd Intercept | 0.51 | | | |
| WAI-S-O: goal | Random | TherapistID | sd Intercept | 0.08 | | | |
| WAI-S-O: goal | Random | Residual | sd Observation | 1.26 | | | |
| WAI-S-O: task | Fixed | | Intercept | 20.28 | .143 | 0.00 | [19.96, 20.59] |
| WAI-S-O: task | Fixed | | CTF condition | 0.88 | .097 | 0.00 | [0.69, 1.07] |
| WAI-S-O: task | Random | PatientID | sd Intercept | 0.35 | | | |
| WAI-S-O: task | Random | TherapistID | sd Intercept | 0.00 | | | |
| WAI-S-O: task | Random | Residual | sd Observation | 1.06 | | | |
| WAI-S-O: bond | Fixed | | Intercept | 20.13 | .145 | 0.00 | [19.80, 20.45] |
| WAI-S-O: bond | Fixed | | CTF condition | 2.19 | .091 | 0.00 | [2.02, 2.37] |
| WAI-S-O: bond | Random | PatientID | sd Intercept | 0.37 | | | |
| WAI-S-O: bond | Random | TherapistID | sd Intercept | 0.00 | | | |
| WAI-S-O: bond | Random | Residual | sd Observation | 1.00 | | | |
| TES: total | Fixed | | Intercept | 25.25 | .247 | 0.00 | [24.72, 25.77] |
| TES: total | Fixed | | CTF condition | 14.48 | .285 | 0.00 | [13.92, 15.04] |
| TES: total | Random | PatientID | sd Intercept | 0.31 | | | |
| TES: total | Random | TherapistID | sd Intercept | 0.26 | | | |
| TES: total | Random | Residual | sd Observation | 3.12 | | | |
| Therapist Credibility | Fixed | | Intercept | 1.73 | .039* | 0.00 | [1.65, 1.82] |
| Therapist Credibility | Fixed | | CTF condition | 0.17 | .026* | 0.00 | [0.12, 0.22] |

| | | | | | | | |
|---|---|---|---|---|---|---|---|
| Therapist Credibility | Random | PatientID | sd Intercept | 0.10 | | | |
| Therapist Credibility | Random | TherapistID | sd Intercept | 0.02 | | | |
| Therapist Credibility | Random | Residual | sd Observation | 0.29 | | | |
| Patient Credibility | Fixed | | Intercept | 3.26 | .083 | 0.00 | [3.07, 3.44] |
| Patient Credibility | Fixed | | CTF condition | -0.01 | .056 | 0.90 | [-0.12, 0.10] |
| Patient Credibility | Random | PatientID | sd Intercept | 0.20 | | | |
| Patient Credibility | Random | TherapistID | sd Intercept | 0.01 | | | |
| Patient Credibility | Random | Residual | sd Observation | 0.61 | | | |
| Patient Improvement: Importance | Fixed | | Intercept | 1.31 | .140 | 0.00 | [0.99, 1.63] |
| Patient Improvement: Importance | Fixed | | CTF condition | 0.04 | .050 | 0.44 | [-0.06, 0.14] |
| Patient Improvement: Importance | Random | PatientID | sd Intercept | 0.38 | | | |
| Patient Improvement: Importance | Random | TherapistID | sd Intercept | 0.00 | | | |
| Patient Improvement: Importance | Random | Residual | sd Observation | 0.55 | | | |
| Patient Improvement: Confidence | Fixed | | Intercept | 2.92 | .082 | 0.00 | [2.74, 3.11] |
| Patient Improvement: Confidence | Fixed | | CTF condition | 0.17 | .044* | 0.00 | [0.09, 0.26] |
| Patient Improvement: Confidence | Random | PatientID | sd Intercept | 0.22 | | | |
| Patient Improvement: Confidence | Random | TherapistID | sd Intercept | 0.00 | | | |
| Patient Improvement: Confidence | Random | Residual | sd Observation | 0.48 | | | |
| Therapist lexical diversity (MTLD) | Fixed | | Intercept | 57.99 | .748 | 0.00 | [56.35, 59.63] |
| Therapist lexical diversity (MTLD) | Fixed | | CTF condition | 10.29 | .581 | 0.00 | [9.15, 11.43] |
| Therapist lexical diversity (MTLD) | Random | PatientID | sd Intercept | 1.71 | | | |
| Therapist lexical diversity (MTLD) | Random | TherapistID | sd Intercept | 0.41 | | | |
| Therapist lexical diversity (MTLD) | Random | Residual | sd Observation | 6.36 | | | |
| Patient lexical diversity (MTLD) | Fixed | | Intercept | 53.51 | .948 | 0.00 | [51.37, 55.65] |
| Patient lexical diversity (MTLD) | Fixed | | CTF condition | 0.06 | .471 | 0.90 | [-0.87, 0.98] |
| Patient lexical diversity (MTLD) | Random | PatientID | sd Intercept | 2.51 | | | |

| | | | | | | | |
|---|---|---|---|---|---|---|---|
| Patient lexical diversity (MTLD) | Random | TherapistID | sd Intercept | 0.00 | | | |
| Patient lexical diversity (MTLD) | Random | Residual | sd Observation | 5.16 | | | |
| Mean words per therapist utterance | Fixed | | Intercept | 21.05 | .706 | 0.00 | [19.55, 22.56] |
| Mean words per therapist utterance | Fixed | | CTF condition | 30.15 | .819 | 0.00 | [28.54, 31.76] |
| Mean words per therapist utterance | Random | PatientID | sd Intercept | 0.90 | | | |
| Mean words per therapist utterance | Random | TherapistID | sd Intercept | 0.67 | | | |
| Mean words per therapist utterance | Random | Residual | sd Observation | 8.97 | | | |
| Mean words per patient utterance | Fixed | | Intercept | 70.92 | 1.396 | 0.00 | [68.17, 73.66] |
| Mean words per patient utterance | Fixed | | CTF condition | 6.77 | 1.961 | 0.00 | [2.92, 10.62] |
| Mean words per patient utterance | Random | PatientID | sd Intercept | 0.00 | | | |
| Mean words per patient utterance | Random | TherapistID | sd Intercept | 0.00 | | | |
| Mean words per patient utterance | Random | Residual | sd Observation | 21.48 | | | |
| Total number of utterances per session | Fixed | | Intercept | 47.53 | 1.321 | 0.00 | [44.66, 50.41] |
| Total number of utterances per session | Fixed | | CTF condition | -15.02 | 1.127 | 0.00 | [-17.23, -12.80] |
| Total number of utterances per session | Random | PatientID | sd Intercept | 2.97 | | | |
| Total number of utterances per session | Random | TherapistID | sd Intercept | 0.00 | | | |
| Total number of utterances per session | Random | Residual | sd Observation | 12.34 | | | |
| Total number of words per session | Fixed | | Intercept | 2,036.52 | 65.885 | 0.00 | [1890.96, 2182.08] |
| Total number of words per session | Fixed | | CTF condition | -125.31 | 47.776 | 0.01 | [-219.19, -31.43] |
| Total number of words per session | Random | PatientID | sd Intercept | 159.62 | | | |
| Total number of words per session | Random | TherapistID | sd Intercept | 0.00 | | | |
| Total number of words per session | Random | Residual | sd Observation | 523.12 | | | |

*Notes: Fixed-effect estimates and variance components from linear mixed-effects models for all primary and secondary outcomes. Intercepts correspond to the low-CTF condition; the "High–Low CTF" coefficient gives the estimated condition difference. Models include random intercepts for patient vignette and therapist persona. P-values are calculated according to the Satterthwaite approach. Abbreviations: MITI, Motivational Interviewing Treatment Integrity Code; WAI-S-O, Working Alliance Inventory-Short Observer form; TES, Therapist Empathy Scale; MTLD, measure of textual lexical diversity; CTF, common therapeutic factors.*

**Supplementary Table ST5**

Reliability of repeated LLM-based ratings.

| Outcome | ICC-1K | *F* | *p* | 95% *CI* |
|---|---|---|---|---|
| MITI: cultivating change talk | 0.87 | 7.71 | < .001 | [0.85, 0.89] |
| MITI: softening sustain talk | 0.93 | 14.17 | < .001 | [0.92, 0.94] |
| MITI: partnership | 0.91 | 11.52 | < .001 | [0.90, 0.92] |
| MITI: empathy | 0.91 | 11.00 | < .001 | [0.90, 0.92] |
| Therapist Credibility | 0.94 | 16.88 | < .001 | [0.93, 0.95] |
| Patient Credibility | 0.88 | 8.50 | < .001 | [0.87, 0.90] |
| Patient Improvement: Importance | 0.93 | 13.62 | < .001 | [0.92, 0.94] |
| Patient Improvement: Confidence | 0.92 | 12.24 | < .001 | [0.91, 0.93] |
| WAI-S-O: total | 0.92 | 12.45 | < .001 | [0.91, 0.93] |
| WAI-S-O: goal | 0.89 | 9.34 | < .001 | [0.88, 0.91] |
| WAI-S-O: task | 0.90 | 9.94 | < .001 | [0.88, 0.91] |
| WAI-S-O: bond | 0.93 | 14.23 | < .001 | [0.92, 0.94] |
| TES: total | 0.99 | 98.33 | < .001 | [0.99, 0.99] |

*Notes: Intraclass correlations (ICCs) for repeated GPT-5 ratings across transcripts (N = 514; 5 repeat ratings per transcript). Raters are considered independent across repetitions and rated sessions. "ICC-1K" reflects reliability of the mean of five ratings. Interpretation: poor < 0.40, fair 0.40-0.59, good 0.6-0.75, excellent > 0.75. Abbreviations: ICC, intraclass correlation coefficient; MITI, Motivational Interviewing Treatment Integrity; WAI-S-O, Working Alliance Inventory-Short Observer form; TES, Therapist Empathy Scale.*

**Supplementary Table ST6**

Human-coded subsample: condition differences.

| **Outcome** | ***M*** | **Median** | ***SD*** | ***IQR*** | ***t*** | ***df*** | ***p*** | ***d*** | **95% *CI*** |
|---|---|---|---|---|---|---|---|---|---|
| MITI: Change Talk | 3.84 | 4 | 0.8 | 1 | 2.93 | 37.25 | < .01 | 0.91 | [0.26, 1.56] |
| MITI: Sustain Talk | 3.59 | 3.5 | 0.73 | 1 | 2.67 | 35.93 | < .05 | 0.83 | [0.18, 1.47] |
| MITI: Partnership | 3.85 | 4 | 0.82 | 1.5 | 4.48 | 35.22 | < .001 | 1.39 | [0.70, 2.08] |
| MITI: Empathy | 3.84 | 4 | 0.76 | 1.12 | 6.28 | 33.15 | < .001 | 1.95 | [1.19, 2.70] |
| Therapist Credibility | 4 | 4 | 1.86 | 2 | -1.46 | 37.92 | .152 | -0.45 | [-1.08, 0.17] |
| Patient Credibility | 5.42 | 5.5 | 2.14 | 3.12 | -0.15 | 36.6 | .885 | -0.05 | [-0.67, 0.57] |
| Patient improvement: Importance | 1.39 | 1 | 1 | 1.12 | -0.39 | 37.92 | .699 | -0.12 | [-0.74, 0.50] |
| Patient improvement: Confidence | 3.41 | 3 | 1.28 | 1 | 0.43 | 37.46 | .671 | 0.13 | [-0.49, 0.75] |
| WAI-S-O: Total | 65.5 | 66.75 | 8.57 | 9.62 | 3.31 | 36.25 | < .01 | 1.02 | [0.36, 1.69] |
| WAI-S-O: Goal | 21.44 | 21.5 | 3.06 | 3.5 | 1.95 | 37 | .059 | 0.6 | [-0.03, 1.24] |
| WAI-S-O: Task | 22.35 | 22.75 | 2.82 | 4.12 | 2.51 | 35.56 | < .05 | 0.78 | [0.14, 1.42] |
| WAI-S-O: Bond | 21.71 | 22 | 3.66 | 3.38 | 4.21 | 35.1 | < .001 | 1.31 | [0.62, 1.99] |
| TES: Total | 40.35 | 41 | 8.15 | 12.25 | 6.46 | 37.33 | < .001 | 2 | [1.24, 2.77] |

*Notes: Descriptive statistics and estimated High–Low CTF differences for MITI, TES, and WAI-S-O outcomes in the independently rated subset. Parametric estimates (Welch's t-test) are shown; nonparametric sensitivity tests yielded the same direction of effects (available on request). Abbreviations: MITI, Motivational Interviewing Treatment Integrity; WAI-S-O, Working Alliance Inventory-Short Observer form; TES, Therapist Empathy Scale*

**Supplementary Table ST7**

Descriptive characteristics of simulated sessions by CTF condition.

| Outcome | *M* | | Median | | *SD* | | *IQR* | |
|---|---|---|---|---|---|---|---|---|
| | low | high | low | high | low | high | low | high |
| MITI: Change Talk | 3.76 | 4.30 | 4.00 | 4.20 | 0.34 | 0.34 | 0.40 | 0.60 |
| MITI: Sustain Talk | 3.16 | 4.11 | 3.20 | 4.20 | 0.63 | 0.38 | 1.00 | 0.20 |
| MITI: Partnership | 3.81 | 4.67 | 4.00 | 4.80 | 0.36 | 0.31 | 0.40 | 0.60 |
| MITI: Empathy | 3.48 | 4.23 | 3.40 | 4.00 | 0.42 | 0.29 | 1.00 | 0.40 |
| WAI: Total | 60.18 | 63.28 | 60.00 | 63.00 | 3.05 | 2.75 | 2.80 | 3.40 |
| WAI: Goal | 19.77 | 19.82 | 19.60 | 19.60 | 1.33 | 1.40 | 1.40 | 1.60 |
| WAI: Task | 20.28 | 21.14 | 20.20 | 21.00 | 1.05 | 1.18 | 0.80 | 1.80 |
| WAI: Bond | 20.13 | 22.32 | 20.40 | 22.40 | 1.27 | 0.83 | 1.60 | 1.20 |
| TES: Total | 25.24 | 39.72 | 25.00 | 39.60 | 4.00 | 1.99 | 5.40 | 2.80 |
| Change in Importance | 1.31 | 1.35 | 1.00 | 1.00 | 0.72 | 0.62 | 0.40 | 0.60 |
| Change in Confidence | 2.92 | 3.09 | 3.00 | 3.00 | 0.52 | 0.53 | 0.60 | 0.60 |
| Therapist Credibility | 1.74 | 1.90 | 1.80 | 2.00 | 0.32 | 0.29 | 0.60 | 0.20 |
| Patient Credibility | 3.26 | 3.25 | 3.20 | 3.20 | 0.71 | 0.58 | 1.00 | 0.80 |
| Total number of utterances | 47.48 | 32.57 | 45.00 | 32.00 | 16.08 | 8.21 | 19.00 | 10.50 |
| Total number of words | 2,032.92 | 1,913.64 | 1,954.00 | 1,904.00 | 636.91 | 444.52 | 793.00 | 580.00 |
| Mean words per therapist utterance | 21.06 | 51.21 | 20.59 | 49.29 | 3.98 | 12.11 | 5.20 | 15.27 |
| Mean words per patient utterance | 70.92 | 77.69 | 68.35 | 73.23 | 19.39 | 23.43 | 26.05 | 32.06 |
| Therapist lexical diversity | 58.03 | 68.25 | 57.94 | 67.82 | 6.98 | 6.23 | 9.80 | 7.68 |
| Patient lexical diversity | 53.53 | 53.53 | 53.47 | 53.04 | 5.82 | 5.68 | 7.33 | 7.03 |

*Notes: Descriptive statistics of simulated sessions for MITI, TES, and WAI-S-O outcomes. Abbreviations: CTF, common therapeutic factors; MITI, Motivational Interviewing Treatment Integrity Code; WAI, Working Alliance Inventory; TES, Therapist Empathy Scale.*
